# Evidence-based blood tests for monitoring adults with chronic kidney disease stage 3 in primary care: rapid review, routine data analysis, and consensus study

**DOI:** 10.1101/2025.04.28.25326552

**Authors:** Martha M C Elwenspoek, Rachel O’Donnell, Catalina Lopez Manzano, Lewis Buss, Sarah Dawson, Katie Charlwood, Thomas A Harding, Christina Stokes, Francesco Palma, Alastair D Hay, Jessica Watson, Penny Whiting

## Abstract

**Main problem:** There is substantial UK variation regarding which blood tests people receive for routine monitoring of stage 3 chronic kidney disease (CKD3) in primary care, suggesting that many people are not receiving optimal care. This study aimed to develop evidence-based testing panels for CKD3.

**Methods:** We considered blood tests used commonly or recommended in CKD3 guidelines, identifying the rationale for each test and then applying a series of filtering questions (e.g. ‘can the GP do anything in response to an abnormal result?’) to identify whether tests should be included in the testing panel. Each question was answered by stepwise rapid evidence reviews. A consensus group, consisting of patient representatives and clinicians, voted on whether tests should be included based on the evidence. If tests had insufficient evidence, additional evidence was collected through rapid reviews or routine data analysis and voted on in a second consensus meeting.

**Results:** There was good evidence for, and consensus supporting, routinely testing eGFR, haemoglobin, and HbA1c; and not routinely testing urea, lipids, vitamin B12, ferritin, folate, liver function, potassium, sodium, vitamin D, calcium, thyroid function, clotting, C-reactive protein, erythrocyte sedimentation rate, and B-type natriuretic peptide. Once on stable treatment, patients on statins do not need additional monitoring tests and patients on ACE-1 or ARB only need potassium monitoring.

**Conclusions:** In contrast to current guidelines, our findings suggest that CKD3 patients should only be routinely offered eGFR, HbA1c and haemoglobin monitoring. Implementing these recommendations could reduce testing variation in CKD3 patients and reduce costs.

**Lay summary:** Patients with chronic kidney disease are often followed up by their GP. At regular monitoring appointments, blood and urine tests are done to determine whether a change in management is needed, such as adjusting medications. However, which tests are needed at these appointments is not standardised and depending on where you live, the tests that you get may differ because of differences in local practices. Here, we reviewed the evidence for blood tests that are currently used for patients with chronic kidney disease, and we found that for many tests there is little or no evidence that they are beneficial. We only found good evidence for regular testing of eGFR to test renal function, HbA1c to detect diabetes, and haemoglobin to detect anaemia. Avoiding unnecessary tests is important to prevent overdiagnosis, overtreatment, patient anxiety, and to ensure that resources are focused on effective, evidence-based care.

## Introduction

In the UK, an estimated 1.9 million adults have a diagnosis of chronic kidney disease (CKD).^1^ CKD is a persistent abnormality in kidney structure or function with type 2 diabetes and hypertension as major risk factors. The prevalence of CKD is expected to continue to increase due to an increasing incidence of type 2 diabetes and hypertension, as well as an aging population.^2,3^ CKD stages 1-2 (CKD1-2) suggest mildly reduced kidney function which does not necessarily require routine monitoring. Patients with CKD3 have impaired kidney function and monitoring and treatment at this stage is important because progressive CKD is associated with adverse clinical outcomes, including end-stage renal disease, cardiovascular disease, and mortality.^4–6^ CKD4-5 represents significant impairment of kidney function which is usually monitored in secondary care and may require dialysis.

CKD3 is primarily managed in primary care. Guidelines suggest that eGFR and urine albumin:creatinine ratio (ACR) should be monitored at least once a year to at least every three months depending on the severity of CKD.^7^ This recommendation is based on a combination of expert opinion and evidence.^8^ However, when asking GPs what they would order routinely for an average patient with CKD3, a substantial proportion of GPs said they would add additional tests, such as full blood count (48%), lipids (38%), HbA1c (23%), and liver function tests (18%).^9^ Similarly, routinely collected UK primary care data show that additional tests are requested and that the most commonly ordered blood tests in people with CKD3 are renal function tests, liver function tests, full blood count, lipids, and HbA1c. Importantly, there is substantial variation in the use of these tests between GP practices, with testing rates varying 40-70% between high and low testing practices, which cannot be explained by variations in demographics of the patient population.^10^

Variation in monitoring suggests that not everyone receives the tests that they need and compared to the guidelines, some people may receive more tests than they need. Underuse of tests risk missing disease progression or new diagnoses and delay treatment. Overuse of tests increases the risk of picking up transient and clinically unimportant fluctuations, borderline results, and false positive results, which lead to unnecessary follow-up tests, appointments, referrals, invasive procedures, and treatments.^11^

Therefore, it is key to optimise the use of monitoring tests to ensure that patients receive the tests that they need without increasing the number of unnecessary tests. To do this, we set out to investigate the rationale of all commonly used blood tests in adults with CKD3, and to identify the evidence that supports their use. We focused on CKD3 because it benefits from monitoring and can be managed in primary care. The aim was to create a minimal set of evidence-based tests that should be offered to all adults with CKD3 and to identify which tests should not be offered routinely.

## Methods

### Identifying candidate tests

We identified a list of ‘candidate’ tests that are commonly used, based on a survey of primary care clinicians^9^ and routinely collected primary care data (CPRD),^10^ or recommended by current UK guidelines.^8^

We determined the potential rationale for ordering each candidate test: detect disease progression, monitor treatment response, or detect secondary complications, and adverse treatment effects in adults with CKD3. For adverse treatment effects, we did not consider the use of monitoring tests during treatment initiation and titration and we limited our investigation to the most common first line treatments (statins, angiotensin-converting enzyme inhibitors [ACE-i], and angiotensin 2 receptor blockers [ARB]) and known side-effects listed by the British National Formulary.

### Filtering questions

We determined a list of filtering questions all of which needed to be answered with ‘yes’ for a test to be a useful monitoring test. For monitoring disease progression or treatment response, these were: 1) Can you intervene to improve treatment response or to slow disease progression? 2) Are there clear benefits of earlier detection or treatment? For secondary condition screening, these were: 1) Are people with CKD3 at an increased risk of developing the secondary condition that the test is able to detect? 2) Is there anything the clinician can do to manage or treat this? 3) Are there clear benefits of earlier detection or treatment? For adverse treatment effects, these were: 1) Are people with CKD3 at an increased risk of developing the side effect that the test is able to detect? 2) Is there anything the clinician can do to manage or treat this? 3) Are there clear benefits of earlier detection or treatment?

### Rapid review methods

We used a stepwise approach to answer each filtering question. We moved to the next step if no evidence or evidence of insufficient quality was identified. We considered a filtering question answered (either positively or negatively) if we identified good evidence to answer the question; we then moved to the next question.

In step 1, we extracted recommendations from NICE guidelines and identified the evidence base for each recommendation. Where recommendations were not evidence-based (i.e. based on expert consensus) we moved to step 2. In step 2, we reviewed systematic reviews and meta-analyses. In step 3, we reviewed primary studies restricting our search to known large and relevant datasets, such as NHANES, and European primary care records, such as CPRD. In step 4, we repeated searches from step 3 without restrictions (Table 1).

**Table 1.** Summary of rapid review methods.

| Step | Sources | Restrictions | Screening |
| --- | --- | --- | --- |
| 1 – NICE guidelines | NICE guidelines | None | Independently by two reviewers |
| 2 – Systematic reviews | KSR evidence* | English language, published >2015 | Independently by two reviewers |
| 3 – Primary studies using known relevant datasets | MEDLINE, Embase, and/or CENTRAL | Known large primary care or representative population samples | 20% independently by two reviewers, conflicts were discussion, remaining 80% by one reviewer following Cochrane rapid review guidance<sup>16</sup> |
| 4 – All primary studies | MEDLINE, Embase, and/or CENTRAL | None | 20% independently by two reviewers, conflicts were discussion, remaining 80% by one reviewer |

\* A database that includes all systematic reviews and meta-analyses in healthcare published since 2015.

**Table 3.**
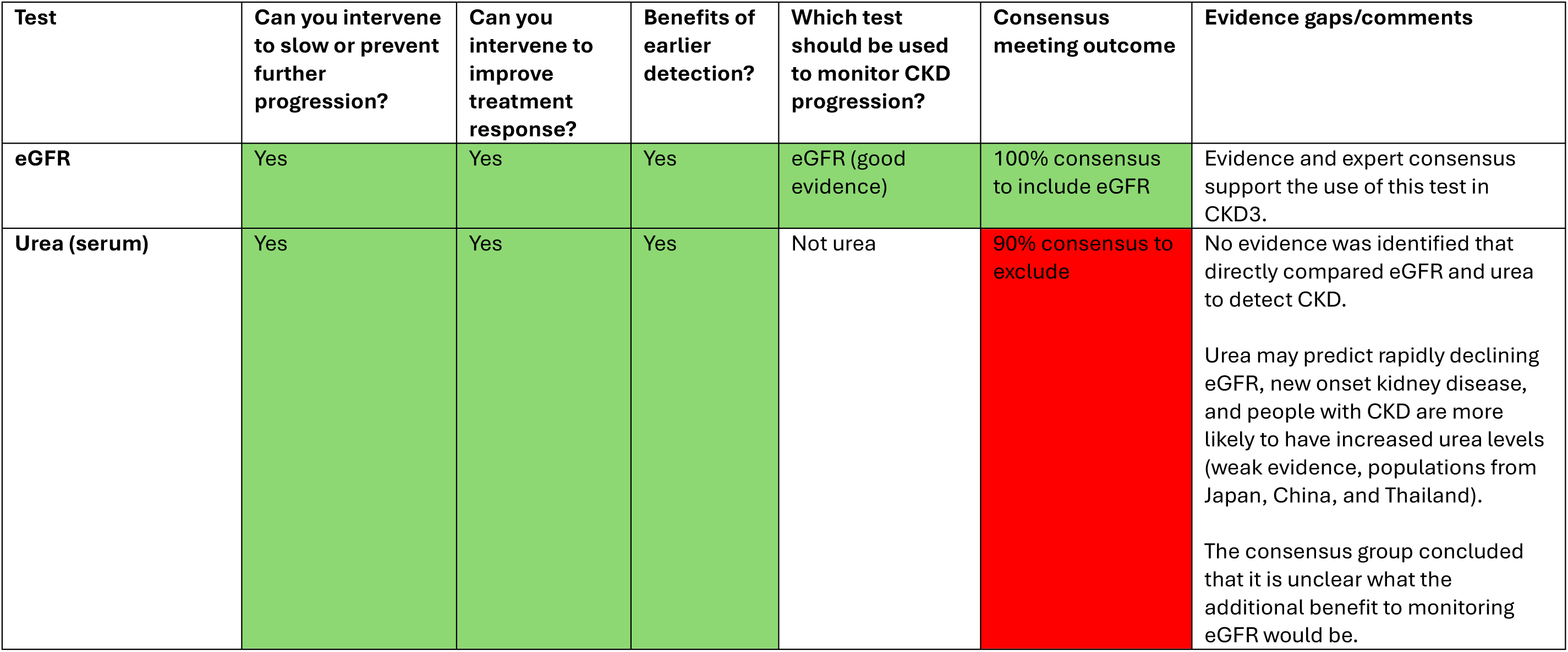
Tests to monitor disease progression and treatment response – answers to filtering questions.

All literature searches were developed by an information specialist (SD) (see Supplementary Tables S1-7 for full search strategies). For each search, we combined terms for CKD with terms for specific tests, secondary conditions, or side effects. Title and abstract screening was performed in Rayyan,^12^ followed by full text screening in Excel. Standardised data extraction forms were developed in Excel and tested by two reviewers and amended where necessary before use. The inclusion criteria, number of included studies and patients, and summary estimates were extracted from included reviews. The study design, population characteristics, number of included patients, and main results were extracted from included primary studies. Risk of bias (ROB) was assessed using ROBIS for systematic reviews,^13^ RoB 2 for RCTs,^14^ and ROBINS-E for cohort studies.^15^ One reviewer conducted data extraction and risk of bias assessments.

### Evidence report and consensus process

We recruited three patient representatives, four GPs, one primary care nurse practitioner, and one renal consultant to form the consensus group. An evidence report summarising all identified evidence was circulated to the consensus group (see Supplementary materials – Evidence report). Evidence was judged as “good” if we found high quality meta-analyses and/or high quality studies with large sample sizes, “moderate” if we found meta-analysis with some quality concerns and/or several primary studies showing similar results but some may have quality concerns, or “weak” if the evidence consisted of a single primary study with a small sample size with or without quality concerns or several single primary studies with conflicting results.

At a meeting, we presented the evidence to the consensus group who voted for each test. Members were asked to vote on whether each candidate test should be included in the panel (i.e. should be offered to all adults with CKD3) or excluded from the panel (should not be offered unless there is an additional clinical indication) or whether there was insufficient evidence to make a recommendation. If no consensus was reached after the first vote (<80%), the group were invited to discuss the evidence, followed by a second vote. If no consensus was reached after two votes, the test progressed to the next stage where additional evidence was generated to help determine whether to include the test.

### Additional evidence and second consensus meeting

In response to specific questions raised by the consensus group, we conducted two additional rapid reviews using the methods described in step 4. The first addressed the question whether monitoring urea provides additional benefit if eGFR is already being monitored. In the second review we searched for an estimate of the incidence of type 2 diabetes in people with CKD.

We also performed analyses in routinely collected primary care records. We used data from patients who were diagnosed with CKD3 and enrolled at a practice contributing to the Clinical Practice Research Datalink (CPRD) GOLD between 2004 and 2019. CPRD Gold is a pseudonymised primary care dataset covering 4% of the UK population,^17^ and is representative of the UK general population in terms of age, sex, and ethnic group.^17^ CPRD records were linked to Hospital Episode Statistics Admitted Patient Care (HES APC),^18^ the Office of National Statistics (ONS) death registration,^19^ the CPRD pregnancy registry, and Patient Level Index of Multiple Deprivation (IMD).

We performed Cox regression comparing people with CKD3 and age-sex- and practice-matched controls to estimate the incidence of abnormal sodium and potassium levels and hypothyroidism in people who had normal levels at diagnosis (methods described elsewhere).^20^ In a second analysis, we aimed to estimate the potential patient benefit of regular monitoring with adjusted calcium. We compared two groups of people with CKD3 in CPRD who attended regular monitoring appointments with and without adjusted calcium testing and followed them for the available follow up time (maximum of 3 years after diagnosis). Patients with a diagnosis of CKD3 with an eGFR within 30 days of diagnosis were identified from CPRD data. We used CPRD data on patient testing within the next 18 months after the initial diagnosis to assign patients into two groups, intervention and control, based on the whether or not they underwent adjusted calcium testing at their next (post diagnostic) eGFR monitoring test. Patients who received an adjusted calcium and eGFR test were assigned to the intervention and patients who only received eGFR were controls. We estimated the incidence of hospitalisations and deaths caused by hyperparathyroidism or bone related conditions (e.g. osteoporosis and fragility fractures), because these conditions can be picked up by adjusted calcium. Code lists for these outcomes were developed with the assistance of two GPs (JW, ADH) with experience working with routine data (Supplementary Table S11).

At a second meeting, the additional evidence and analyses were presented to the consensus group and voted on as described above. The second consensus group consisted of three patient representatives, five GPs, two primary care nurse practitioners, and one renal consultant. If no consensus was reached after two rounds of voting, we asked the group what their recommendation would be (“include” or “exclude”) in absence of any further evidence.

### Patient and Public Involvement

Patient representatives CS and FP were involved in this study from inception. They reviewed the grant application and this manuscript, in particular the plain language summary, and were active members of the project management group who met monthly. Three patient representatives were part of the consensus group with the same voting rights as other group members.

## Results

### Candidate tests

Twenty-one candidate tests were considered. Tables 4-6 summarise the answers to the filtering questions for each of the tests. A detailed evidence report can be found in the Supplementary Materials.

**Table 4.** Tests to screen for secondary conditions – answers to filtering questions.

| Test | Reason for testing | Are people with CKD3 at risk? | Can you intervene? | Benefits of earlier detection? | Consensus meeting outcome | Offer routinely to everyone with CKD3? | Evidence gaps/comments |
| --- | --- | --- | --- | --- | --- | --- | --- |
| **Haemoglobin (full blood count)** | To detect anaemia | Yes (moderate evidence) | Yes | Yes | 100% consensus to include | YES | Evidence and expert consensus support the use of this test in CKD3. |
| **HbA1C** | To detect T2DM | Yes (moderate evidence) | Yes | Yes | 90% consensus to include | YES | People with CKD have a higher incidence of T2DM than in the general population (moderate evidence) and lower levels of eGFR are associated with an increased risk of new onset T2DM (moderate evidence). |
| **Potassium (electrolytes)** | To detect hyper- or hypokalaemia | Yes (weak evidence) | Yes | Yes | 100% consensus to exclude | NO | The advisory board agreed after three rounds of voting that this test should be excluded for patients who are not on any medications and <CKD4. Arguments against monitoring were that chronically elevated potassium levels are uncommon, acute dramatic increases are clinically relevant but will not be picked up by annual monitoring, and blood samples are not processed quickly enough which causes false positives. |
| **Sodium (electrolytes)** | To detect hyper- or hyponatraemia | No (weak evidence) | Yes | Yes | 100% consensus to exclude | NO | Incidence analyses showed that clinically significant abnormal sodium levels were |
|  |  |  |  |  |  |  | NOT more common in CKD than in the control group. However, this test will get added automatically to eGFR in many labs. |
| **Vitamin D** | To detect vitamin D deficiency | Yes (weak evidence) | Yes | Unclear | 90% consensus to exclude | NO | The rationale for testing was questioned, as vitamin D supplementation is recommended to everyone in the UK during winter and autumn,<sup>32</sup> it may not give any helpful information for CKD management. |
| **Adjusted calcium (bone profile)** | To detect hypo- or hypercalcaemia | No (hypocalcaemia, moderate evidence; hypercalcaemia, no evidence) | Yes | Unclear | 80% consensus to exclude | NO | We found no hospitalisations caused by hyperparathyroidism or bone related conditions (e.g. osteoporosis / fragility fractures) in the first three years after CKD3 diagnosis suggesting that these events are extremely rare. However, it is unclear if this changes beyond three years. |
| **Thyroid function tests** | To detect hyper- or hypothyroidism | Yes (hyperthyroidism, no evidence; hypothyroidism, moderate evidence) | Yes | Unclear | 88% consensus to exclude | NO | The incidence of hypothyroidism in patients with CKD who had normal thyroid function at diagnosis is similar to that of the control group. |
| **Lipid profile** | To detect dyslipidaemia, | Yes (dyslipidaemia, weak evidence; | Yes | No | 88% consensus to exclude | NO | Everyone with CKD are prescribed statins regardless |
|  | assess cardiovascular disease risk | cardiovascular risk, good evidence) |  |  |  |  | of lipid levels to reduce cardiovascular risk. The consensus group believed testing could do more harm than good by dissuading patients to take statins (or follow lifestyle advice) if their lipids are normal. |
| **Ferritin** | To detect ferritin deficiencies and related anaemias | Yes (weak evidence) | Yes | Yes | 86% consensus to exclude | NO | The consensus group decided that ferritin is not required in addition to haemoglobin as this will detect anaemia. |
| **Folate** | To detect folate deficiencies and related anaemias | Unclear (no evidence) | Yes | Yes | 86% consensus to exclude | NO | The consensus group decided that folate is not required in addition to haemoglobin as this will detect anaemia. |
| **Vitamin B12** | To detect vitamin B12 deficiencies and related anaemias | Unclear (no evidence) | Yes | Yes | 86% consensus to exclude | NO | The consensus group decided that vitamin B12 is not required in addition to haemoglobin as this will detect anaemia. |
| **Liver function tests** | To detect metabolic dysfunction-associated steatotic liver disease (MASLD) | No (moderate evidence) | No* | Yes | 100% consensus to exclude | NO | The consensus group agreed that monitoring liver function did not benefit patient outcomes due to the lack of treatments for MASLD. |
| **Clotting tests** | To detect bleeding or clotting disorders | Unclear (no evidence) |  |  | 100% consensus to exclude | NO | No evidence was identified on the risk of bleeding disorders amongst CKD patients. |
| **C-reactive protein (CRP) and erythrocyte sedimentation rate (ESR)** | General marker for inflammation (and infection), but are not linked to any particular condition | Yes (good evidence) | No (only lifestyle advice) | Unclear | 100% consensus to exclude | NO | There is no guidance on how to treat low grade inflammation. It is unclear whether reducing CRP or ESR directly has any benefit. |
| **B-type natriuretic peptide (BNP)** | To detect heart failure | Yes (weak evidence) |  |  | 88% consensus to exclude | NO | It was noted that this test has low accuracy for heart failure and was not developed as a monitoring test. |

\*This may change in the future if new medications become available.
Abbreviations: BNP, B-type natriuretic peptide; CKD, chronic kidney disease; CRP, C-reactive protein ; ESR, erythrocyte sedimentation rate; MASLD, metabolic dysfunction-associated steatotic liver disease; T2DM, type 2 diabetes mellitus;

**Table 5.** Tests to screen for adverse treatment effects of statins and ACE-i/ARBs for patients on stable treatment – answers to filtering questions.

| Test | Purpose of test | Are people on these medications at risk? | Can you intervene? | Benefits of earlier detection? | Consensus meeting outcome | Offer routinely to everyone on these medications? | Evidence gaps/comments |
| --- | --- | --- | --- | --- | --- | --- | --- |
| **Tests to screen for adverse effects of statins** |  |  |  |  |  |  |  |
| **Anaemia tests** (including haemoglobin, iron, folate, and vitamin B12) | To detect anaemia, iron deficiency, folate deficiency and vitamin B12 deficiency | No (not reported by BNF) | Yes (change medications) | Yes (change medications sooner) | 88% consensus to exclude this test | NO | No evidence suggesting that there is an increased risk of anaemia or deficiencies associated with statin use. |
| **Renal function tests** (including eGFR and blood electrolytes) | To detect CKD, hyper- or hypokalaemia, hyper- or hyponatremia | No (not reported by BNF) | Yes (change medications) | Yes (change medications sooner) | 88% consensus to exclude this test | NO | The exception to this is Rosuvastatin, however, this is not commonly prescribed and so routine monitoring of all patients on statins would not be appropriate. |
| **Liver function tests** | To detect liver disease and liver damage | No (not reported by BNF) | Yes (change medications) | Yes (change medications sooner) | 88% consensus to exclude this test | NO | Changes to liver profile in general is considered a drug class effect and liver function would be tested when first starting a medication and so regular monitoring would not be needed. |
| **HbA1c** | To detect new onset diabetes | No (not reported by BNF) | Yes (change medications) | Yes (change medications sooner) | 100% consensus to exclude this test | NO | No evidence to suggest that statins are associated with a greater risk of new-onset diabetes. |
| **Tests to screen for adverse effects of ACE-i/ARBs** |  |  |  |  |  |  |  |
| **Anaemia tests** | To detect anaemia, iron deficiency, folate acid deficiency and vitamin B12 deficiency | Unclear (no evidence) | Yes (change medications) | Yes (change medications sooner) | 90% consensus to exclude | NO | The BNF lists anaemia as rare (1 in 10 000 to 1 in 1000) or very rare (<1 in 10 000) adverse events but does not cite evidence. |
| **eGFR** | To detect kidney injury | Unclear (inconsistent evidence) | Yes (change medications) | Yes (change medications sooner) | 80% consensus to exclude | NO | ACE-i/ARBs preserve renal function by making them work slower, which is reflected in an initial drop in renal function which is renal protective in the long term. Renal function will be monitored as part of the panel. |
| **Potassium** | To detect hyper- or hypokalaemia | Yes (good evidence) | Yes (change medications) | Yes (change medications sooner) | 80% consensus to include | YES | There is good evidence that ACE-I and ARBs increase the risk of hyperkalaemia. |
| **Liver function tests** | To detect liver disease and liver damage | Unclear (no evidence) | Yes (change medications) | Yes (change medications sooner) | 100% consensus to exclude | NO | The British National Formulary (BNF) lists hepatic disorders, hepatitis, or abnormal hepatic function as rare (1 in 10 000 to 1 in 1000) or very rare (<1 in 10 000) adverse events but does not cite evidence. |
| **HbA1c** | To detect new onset T2DM | Unclear (no evidence) | Yes (change medications) | Yes (change medications sooner) | 89% consensus to exclude | NO | ACE-i and ARB reduce the risk to develop T2DM compared to placebo (good evidence). |

### Tests with good evidence and rationale

We identified good evidence supporting the use of eGFR to monitor disease progression and treatment response and haemoglobin (as part of full blood count) to detect anaemia. Both tests were included in the final panel with 100% consensus at the first consensus meeting. The evidence for using HbA1c to detect type 2 diabetes in people with CKD3 was insufficient and no consensus was reached at the first meeting. The consensus group requested further evidence on the incidence of new onset type 2 diabetes in adults with CKD3. We identified moderate level evidence suggesting that new onset type 2 diabetes is indeed higher in adults with CKD3.^21–25^ As a result, there was 90% consensus to include HbA1c in the testing panel (Tables 4-5).

### Tests with unclear evidence or rationale

We identified weak evidence showing that the prevalence of hyperkalaemia increases as renal function declines.^26,27^ Additional analyses in CPRD suggested that clinically significant abnormal potassium levels were more common in CKD3 than in the control group (moderate evidence).^20^ However, this analysis did not control for medication use, which may be the reason for the increased incidence of abnormal potassium levels. After three rounds of voting and discussions, the consensus group agreed that there was insufficient evidence to justify regular testing of sodium and potassium in CKD3 patients who are not on specific medications, such as ACE-I or ARBs. However, many CKD3 patients are using these medications and most laboratories will measure sodium and potassium together with eGFR, so this recommendation may have limited impact in practice.

Although we identified a small case control study and a large cross sectional survey that suggest that the prevalence of overt and subclinical hypothyroidism increases as eGFR declines,^28,29^ it is unclear whether patients with normal thyroid function (test results within the normal/reference range) at diagnosis are more likely to develop hypothyroidism later, suggesting that one measurement at CKD diagnosis is likely to be sufficient (Table 5). The consensus panel agreed to exclude thyroid tests from the panel in absence of more evidence.

We identified moderate evidence that the prevalence of hypocalcaemia is similar in people without CKD and people with CKD1-3 but increases as CKD progresses to CKD4.^30,31^ It is unclear if CKD is associated with hypercalcaemia (no evidence). We found no hospitalisations caused by hyperparathyroidism or bone related conditions (e.g. osteoporosis or fragility fractures) in the first three years after CKD3 diagnosis (n=19,554) in routine data (moderate evidence). These findings suggest that monitoring adjusted calcium may not be beneficial in the first 3 years after diagnosis, since these events are extremely rare. The consensus panel agreed to exclude calcium tests from the panel in absence of more evidence.

### Tests that should not be routinely offered to patients with CKD3

The evidence and consensus meeting results suggest that the following tests should not be offered routinely to patients with CKD3, unless there are clear additional clinical indications (Table 4-6). Urea should not replace eGFR to monitor CKD progression. There is no need to offer regular monitoring of vitamin D, lipid profile, ferritin, folate, vitamin B12, liver function, clotting, c-reactive protein (CRP), erythrocyte sedimentation rate (ESR), or B-type natriuretic peptide (BNP) to detect secondary conditions. Changes to liver function in general is considered a drug class effect and liver function would be tested when first starting a medication; however, subsequent monitoring of liver function would not be necessary.

## Discussion

### Summary

There is good evidence to support monitoring eGFR, haemoglobin, and type 2 diabetes in all patients with CKD3. We identified evidence suggesting that routine use of the following tests are not beneficial for patients and should not be offered as part of regular monitoring appointments: urea, vitamin D, thyroid function, lipid profile, ferritin, folate, vitamin B12, liver function, clotting, CRP, ESR, or BNP. The evidence base for sodium, potassium, and calcium for CKD3 is either weak or absent and there was consensus that these tests should not be offered in absence of further evidence. Once on stable treatment, we found no evidence for the need of additional monitoring tests for patients on statins and for patients on ACE-1 or ARB we only found evidence in support of potassium monitoring.

### Strengths and limitations

A strength of this study is that we have used a standardised approach to identify evidence using established evidence synthesis methods. Evidence gaps identified by the consensus group were addressed with additional systematic literature searches as well as analyses of routinely collected primary care data. We included a diverse consensus group including patients to vote on tests when the evidence was less clear or to give additional context. We also involved patients throughout the project. Final decisions were made by the consensus group. Where expert opinion was used, we documented the rationale for these decisions.

A limitation is that we needed to make compromises in evidence synthesis methods to make the volume of rapid reviews manageable. For instance, we used a stepwise approach where we stopped screening once good evidence was identified, so there is a possibility that we may have missed relevant evidence. We also did not include ACR or other urine tests as there is already clear and evidence-based guidance on ACR monitoring,^7^ and we focussed on CKD3 because most people monitored in primary care have CKD3. Different tests may be necessary as CKD progresses. Another important limitation is that we focussed on commonly used tests and did not include new tests or new treatments. However, these methods can be used to assess the evidence and rationale of any new tests for CKD3. We also did not take test accuracy or biological variation into account, which can play a role in the usefulness of a monitoring test. Due to the lack of evidence, it was not possible to base all decisions on evidence alone, so the final minimal panel is based both on evidence and expert opinion. Finally, although we aimed to include diverse and representative members in the consensus group, it is possible that their views might not have been fully representative of wider patient and clinical communities.

### Comparison with existing literature

A survey among UK GPs showed that eGFR, full blood count, lipid profile, HbA1c, and liver function were among the most commonly ordered tests for ‘an average patient with CKD3’.^9^ Substantial proportions of GPs said to offer these tests routinely: 48% for FBC, 38% lipid profile, 23% for HbA1c, and 18% for liver function tests.^9^ Similarly, routine data from UK primary care showed that renal function tests (2 tests per person-year), liver function tests (1.3 tests per person-year), full blood count (1.3 test per person-year), lipid profile (0.8 tests per person-year), and HbA1c (6.5 tests per person-year) were the most commonly ordered tests for patients with CKD3.^10^ Although liver function tests and lipid profile are commonly ordered in primary care to monitor people with CKD, our results suggest that this is not supported by evidence. However, many people with CKD are prescribed statins and recent guidelines recommend clinicians should ‘consider’ annual lipid monitoring for people on statins, which could explain these high numbers.^33^ The word ‘consider’ is used in NICE guidelines if there is no strong evidence base for the recommendation and the benefit of the recommendation is less certain.

Our findings partly align with NICE guidance.^7^ Similar to our findings, NICE recommends regular monitoring of eGFR and to not offer routine monitoring of vitamin D and calcium to patients with CKD1-3. In contrast to our minimal panel, monitoring haemoglobin and HbA1c are not mentioned in NICE guidelines for CKD3, even though we identified a strong rationale underpinned by evidence to offer these tests routinely to all patients with CKD3. We do not recommend regular lipid testing, whereas NICE recommends treating to target for primary prevention and to aim for >40% reduction in non-HDL cholesterol which suggests that regular monitoring of lipids for people on statins is necessary.^33^ However, statins are believed to be beneficial for all CKD patients to reduce cardiovascular risk, whether they have high cholesterol or not.^7^ As such, routine cholesterol monitoring may have unintended consequences, as individuals could interpret a ‘normal’ result as justification to discontinue their medication.

Finally, a recent HTA report has questioned the benefit of regular monitoring for patients with CKD.^34^ They found that although testing rates have increased dramatically over the years, it is not clear how further decline should be treated, as renoprotective treatments are often already prescribed (such as statins or antihypertensives). They found that eGFR monitoring to guide cardiovascular prevention was not cost-effectiveness mainly because changes in eGFR did not trigger changes in treatment recommendations. The differences between our findings and those of the HTA report are likely due to the fact that our analysis did not take cost-effectiveness into account.

### Implications for research and practice

Clinicians should avoid offering tests routinely in the absence of evidence of benefit, as this can be harmful to patients because it can lead to unnecessary follow up testing, patient anxiety, overdiagnosis, overtreatment, as well as wasting NHS resources. However, testing is often driven by local practice protocols and established habits rather than a specific clinical rationale for each individual case. To change testing practices, local practice protocols need to be adjusted, but also, GP practice staff involved in testing need to be educated on the harms of including unnecessary tests to avoid additional tests to be added back in due to habit. The minimal testing panel that we present here could potentially be implemented in practice using single click buttons in test ordering software, which could address some of the unwarranted variation in testing.

These methods are transferable to other long-term conditions and can be applied to more conditions in the future. The testing panel will need updating when new tests or new treatments become available, such a better treatments for MASLD. Further research into CKD4-5 monitoring is also needed. Finally, there is little evidence on the optimal frequency of testing which needs to be addressed in future research. Methodologically stronger methods (e.g. RCTs) are still needed to establish the safety, effectiveness and cost-effectiveness of adopting this approach.

## Conclusions

In contrast to current guidelines, our findings suggest that CKD3 patients should only be routinely offered eGFR, HbA1c and haemoglobin monitoring. Implementing these recommendations could reduce testing variation and reduce costs.

## Data Availability

The data from rapid reviews and the consensus meetings that support the findings of this study are available within this paper and supplementary files. A small part of this study is based on data from the Clinical Practice Research Datalink (CPRD) obtained under licence from the MHRA and NIHR. The data are provided by patients and collected by the NHS as part of their care and support. CPRD data can be accessed via the MHRA: https://www.cprd.com/

## Authorship and contributions

Martha MC Elwenspoek drafted the manuscript and developed the protocol. Rachel O’Donnell and Catalina Lopez Manzano conducted the abstract screening, full-text screening, data extraction, risk of bias assessment, and wrote the evidence report. Sarah Dawson was responsible for developing all search strategies. Lewis Buss, Tom Harding, and Katie Charlwood conducted CPRD analyses. Alastair Hay and Jessica Watson contributed clinical expertise and secured funding for the project. Penny Whiting developed the methods, secured funding, and provided project oversight together with Jess Watson. All authors had full access to the data in the study and can take responsibility for the integrity and accuracy of the data analysis. All authors have read and approved the final manuscript.

## Ethics approval

For the part of the study that used routine data, the authors were provided with pseudonymised Clinical Practice Research Datalink (CPRD) data under licence from the MHRA and NIHR. The protocol (21_001671) for this study was approved on 21/12/2021 by the Independent Scientific Advisory Committee (ISAC), the independent body that approves use of CPRD data. All data used in this study are routinely collected and anonymised and thus consent was not required. CPRD have approval to collect and disseminate anonymised data to approved researchers for the benefit of public health under IRAS 242149.

## Acknowledgments

We would like to express our gratitude to Aisling O’Rourke for her invaluable administrative support throughout the project and for organising the consensus meeting. We also extend our sincere thanks to the members of the consensus group, including patient representatives Bob Cottis, Jean Palmer, and Patricia MacCalla; GPs Katherine Alsop, Sam Merriel, Rachel Johnson, and Alan McFarlane; renal consultant Dominic Taylor; and primary care nurse Jennifer Charlewood.

Second consensus meeting: patient representatives Bob Cottis, Jean Palmer, and Patricia MacCalla; GPs David Spitzer, Nazmul Mohsin, Tim Johnson, Sam Merriel, and Alan McFarlane; renal consultant Dominic Taylor; and primary care nurses Jennifer Charlewood and Helen Lane.

Additionally, we wish to acknowledge the contributions of the wider team, including Mary Ward, Howard Thom, Alice Malpass, Jonathan Banks, Clare Thomas, Hayley Jones, and Jonathan Sterne.

