## Supplementary data for "Evidence-based blood tests for monitoring adults with chronic kidney disease stage 3 in primary care: rapid review, routine data analysis, and consensus study"

#### Authors

Martha M C Elwenspoek,[1,2] PhD, Rachel O'Donnell,[2] MSc, Catalina Lopez Manzano,[2] MSc, Lewis Buss,[2] MSc, Sarah Dawson,[1] MSc, Katie Charlwood,[2] MSc, Thomas A Harding,[2] MSc, Christina Stokes,[3] Francesco Palma,[3] Alastair D Hay,[2] MD, Jessica Watson,[2] PhD, Penny Whiting,[2] PhD.

#### Affiliations

1. The National Institute for Health Research Applied Research Collaboration West (NIHR ARC West), University Hospitals Bristol NHS Foundation Trust, BS1 2NT Bristol, UK
2. Population Health Sciences, Bristol Medical School, University of Bristol, BS8 2PS Bristol, UK
3. Patient representative

### Search strategies systematic reviews

#### Table S1. Rapid review 1a: Which test should be used to monitor chronic kidney disease?

**Database: KSR Evidence (2015-2021)**

Search for SRs on accuracy of blood urea testing compared to eGFR or accuracy of eGFR or blood urea testing alone

| # | Terms | Hits |
|---|---|---|
| 1 | (CKD or "kidney function*" or "kidney failure*" or "renal function*" or "renal failure*" or nephropathy or nephropathies or nephrologic* or glomerular filtration rate*" or (kidney AND (disorder* or disease* or dysfunction* or chronic* or insufficienc* or impair*)) or (renal AND (disorder* or disease* or dysfunction* chronic* or insufficienc* or impair*)) or ("kidney failure*" NOT (graft* or allograft* or transplant*))) NOT ((renal NEAR (dialysis or surgery or haemodialysis or hemodialysis or transplant)) OR (kidney NEAR (dialysis or surgery or h?emodialysis or transplant))) | 5596 |
| 2 | (analys* or monitor* or test* or manag* or screen* or diagnos* or routine or check* or maintain* or control*) | 210103 |
| 3 | #1 AND #2 | 5280 |
| 4 | eGFR or GFR or cGFR or "Creatinine Clearance" or "Glomerular Filtration Rate" or (GFR and (estimated or calculated)) | 1755 |
| 5 | "Blood urea nitrogen" or (urea and (blood or nitrogen*)) or BUN or (BUN and (level* or test* or serum)) or "urea nitrogen test" | 262 |
| 6 | #3 AND #4 | 783 |
| 7 | #3 AND #5 | 147 |

#### Table S2. Rapid review 2a: How prevalent are the following acute or chronic complications chronic kidney disease stage 3 patients compared to general population?

Including: Liver disease, heart failure, Type 2 diabetes, CVD, anaemia, haeminatics, bone profile related conditions, thyroid disease, and bleeding disorders.

**Database: KSR Evidence**

| # | Terms | Hits |
|---|---|---|
| 1<br>Chronic kidney disease | CKD or “kidney function*” or “kidney failure*” or “renal function*” or “renal failure*” or nephropathy or nephropathies or nephrologic* or glomerular filtration rate*” or (kidney AND (disorder* or disease* or dysfunction* or chronic* or insufficienc* or impair*)) or (renal AND (disorder* or disease* or dysfunction* chronic* or insufficienc* or impair*)) or (“kidney failure*” NOT (graft* or allograft* or transplant*)) not (renal NEAR (dialysis or surgery or haemodialysis or hemodialysis or transplant)) not (kidney NEAR (dialysis or surgery or haemodialysis or hemodialysis or transplant)) | 6596 |
| 2<br>Prevalence | prevalence or incidence or predict* or risk* | 124284 |
| 3<br>Liver disease | “NAFLD” or “liver function” or “fatty liver” or “cirrhosis” or “cirrho*” or “steatohepatitis” or “fibrosis” or “fibro*” or “non-alcoholic | 8771 |

| | | |
|---|---|---|
| | steatohepatitis" or "NASH" or "hepatic*" or "hepato*" or ("hepatic" AND ("complications" or "co-morbid*")) | |
| 4 | #1 AND #3 | 491 |
| 5 | #2 AND #4 | 342 |
| 6<br>Heart failure | "Heart failure" or "cardiac failure" | 3601 |
| 7 | #1 AND #6 | 542 |
| 8 | #2 AND #7 | 455 |
| 9<br>Type 2 diabetes | ("diabetes type 2" or "diabetes type II" or "type 2 diabet*" or "type II diabet*" or (diabet* near ("type 2" or "type II")) or "diabetes mellitus type 2" or "diabetes mellitus type II" or T2DM or "T2 DM" or DMT2 or "DM T2" or "adult onset diabet*" or "late onset diabet*" or "matur* onset diabet*" or "NIDDM" or "non insulin* diabet*" or "noninsulin* diabet*" or "insulin independent diabet*" or "ketosis resistant diabet*") | 5479 |
| 10 | #1 AND #9 | 544 |
| 11 | #2 AND #10 | 451 |
| 12<br><br>Bleeding disorders | (bleeding or platelet AND (disorder* or problem*)) or "platelet function defects" or "Disseminated intravascular coagulation" or DIC or "Prothrombin deficiency" or h?emophilia or "Glanzmann disease" or "Idiopathic thrombocytopenic purpura" or ITP or "Von Willebrand disease" or (factor AND deficiency) | 6891 |
| 13 | #1 AND #12 | 489 |
| 14 | #2 AND #13 | 412 |
| 15 Anaemia (blood count and Haematinics) | "anaem*" or "anem*" or "iron-poor blood" or "Low blood" or "Tired blood" or "iron" or "ferritin" or "folate" or "folic acid" or "vitamin B9" or "B12" or "cobalamin" or "deficien*" or "total iron" or "iron level*" or "levels of iron" or "iron store*" or "stored iron" or "heme iron" or "haem iron" or (("vitamin" or "iron" or "ferritin" or "folate" or "folic acid" or "vitamin B9" or "B12" or "cobalamin") AND ("deficien*" or "low" or "insufficien*" or "lack" or "levels")) | 6427 |
| 16 | #1 AND #15 | 487 |
| 17 | #2 AND #16 | 364 |
| 18 bone profile | "parathyroid*" or "hypoparathyroidism" or "hyperparathyroidism" or "PTH" or "parathormone" or "parathyrin" or "Pseudohypoparathyroidism" or ((bone or bones or 3artilage* or chondro* or osteo* or spine or spinal or skeletal or extraskeletal or Ewing) AND (cancer* or carcinoma* or neoplas* or metast* or sarcoma* or tumo?r*)) or chondrosarcoma* or "vitamin D" or "vitamin D deficien*" or "insufficien* vitamin D" or "Cholecalciferol" or "ergocalciferol" or (("vitamin D" ) AND ("deficien*" or "low" or "insufficien*" or "lack" or "levels")) | 4355 |
| 19 | #1 AND #18 | 299 |
| 20 | #2 AND #19 | 208 |
| 21 thyroid disorders | "Thyroid disease" or "thyroid disorders" or "thyroid dysfunction" or "dysfunctional thyroid" or "hypothyroidism" or "underactive thyroid" or "hyperthyroidism" or "hyper-thyroid*" or "overactive thyroid" or "Hashimoto's" or "Hashimotos" or "thyroiditis" or "Graves'" or "Graves" or "thyroxin*" or "thyroid stimulating hormone" | 925 |
| 22 | #1 AND #21 | 47 |
| 23 | #2 AND #22 | 38 |
| 24<br>Cardiovascular disease | "myocardial infarction" or stroke* or "brain ischemia" or "cerebrovascular accident" or (cardiac NEAR (death or sudden or mortality)) or apoplex* or "Abnormal heart rhythm*" or arrhythmia* or "marfan syndrome" or " Deep vein thrombosis" or "pulmonary | 72717 |

| | | |
|---|---|---|
| | embolism” or “heart attack” or cardiomyopathy or “intracranial h?emorrhage*” or “coronary syndrome” or (cardiovascular NEAR (event* or mortality or death*)) or (disease* or disorder* NEAR (cardiovascular or coronary or heart or artery or cardiac or aorta or “Congenital heart” or “Coronary artery” or “Heart muscle” or “Heart valve” or pericardial or “Peripheral vascular” or “Rheumatic heart” or Vascular or “blood vessel” or “isch?emic heart” or cerebrovascular)) | |
| 25 | #1 AND #24 | 5178 |
| 26 | #2 AND #25 | 3879 |
| | 2020 cut off for CVD | 1656 |

### Table S3. Rapid review 3a: How prevalent are abnormal test results in chronic kidney disease stage 3 patients compared to the general population?

Including: Liver disease tests, lipid tests, full blood count, thyroid function tests, haeminatics, clotting tests, Natriuretic Peptide Tests, bone profile tests, HbA1C

**Database: KSR Evidence**

| # | Terms | Hits |
|---|---|---|
| 1 Chronic kidney disease | (CKD or “kidney function*” or “kidney failure*” or “renal function*” or “renal failure*” or nephropathy or nephropathies or nephrologic* or glomerular filtration rate*” or (kidney AND (disorder* or disease* or dysfunction* or chronic* or insufficienc* or impair*)) or (renal AND (disorder* or disease* or dysfunction* chronic* or insufficienc* or impair*)) or (“kidney failure*” NOT (graft* or allograft* or transplant*))) NOT ((renal NEAR (dialysis or surgery or haemodialysis or hemodialysis or transplant)) OR (kidney NEAR (dialysis or surgery or h?emodialysis or transplant))) | 5384 |
| 2 Prevalence | Prevalence or prevalence studies or incidence or prediction or predict* or risk* | 127177 |
| 3 Liver disease tests | “liver function test*” or “Albumin” or “Alkaline phosphatase” or “ALP” or “Alanine aminotransferase” or “Alanine transaminase” or “ALT” or “Aspartate aminotransferase” or “Aspartate transaminase” or “AST” or “Bilirubin” or “Gamma glutamyl transpeptidase” or “Gamma-glutamyltransferase” or “GGT” or “Total protein” or “Serum globulin” or “Liver enzymes” or “Albumin and total protein” or “L-lactate dehydrogenase” or “LD” or “Prothrombin time” or “PT” | 3082 |
| 4 | #1 AND #3 | 406 |
| 5 | #2 AND #4 | 277 |
| 6 Natriuretic Peptide Tests | “Natriuretic Peptide Tests” or “BNP” or “NT-proBNP” or “brain natriuretic peptide” or “N-terminal pro b-type natriuretic peptide” or “B-type natriuretic peptide” or “brain natriuretic factor” or “BNF” | 434 |
| 7 | #1 AND #6 | 38 |
| 8 | #2 AND #7 | 33 |
| 9 HbA1C | HbA1c or “h?emoglobin A1” or “glycated h?emoglobin” or “glycated ha?emoglobin” or “glycosylated Hb” or GHb or “plasma glucose” or “total A1” or glycoh?emoglobin or HgbA1c or Hb1c | 2606 |
| 10 | #1 AND #9 | 194 |
| 11 | #2 AND #10 | 142 |

| | | |
|---|---|---|
| 12 Clotting tests | "Clotting test*" or "Coagulation Test*" or "Prothrombin time" or "Partial thromboplastin time" or "Thrombin time" or INR or "Factor V assay" or "Fibrinogen level" or "PT" or "PT-INR" or "Platelet count" | 935 |
| 13 | #1 AND #12 | 43 |
| 14 | #2 AND #13 | 36 |
| 15 Anaemia (full blood count) | "FBC" or "full blood count" or "complete blood count" or "CBC" or "Eosinophil count" or "Haemoglobin" or "Mean corpuscular haemoglobin" or "Mean corpuscular haemoglobin concentration" or "Mean corpuscular volume" or "Monocyte count" or "Neutrophil count" or "All platelet tests" or "Red blood cell count" or "Total white blood cell count" or "Lymphocyte count" or "Packed cell volume" or "Basophil count" or "Differential white cell count" or "Red blood cell size" | 1535 |
| 16 | #1 AND #15 | 112 |
| 17 | #2 AND #16 | 100 |
| 18 bone profile | "Bone profile" or "Serum inorganic phosphate" or Calcium or "Bone studies" or "Alkaline phosphatase" | 1894 |
| 19 | #1 AND #18 | 190 |
| 20 | #2 AND #19 | 139 |
| 21 thyroid function tests | "Thyroxine or "T4" or "Thyroid stimulating hormone" or "TSH" or "Thyroid function" or "tri-iodothyronine" or "T3" | 564 |
| 22 | #1 AND #21 | 21 |
| 23 | #2 AND #22 | 14 |
| 24 Lipid tests | "Serum cholesterol" or "total cholesterol" or" High density lipoprotein" or HDL or "Low density lipoprotein" or LDL or Triglyceride* or "HDL:LDL ratio" or "HDL LDL ratio" or "Lipid Profile" or "Fasting Lipid*" or "Non fasting Lipid*" or "Cholesterol Test*" | 3326 |
| 25 | #1 AND #24 | 178 |
| 26 | #2 AND #25 | 124 |
| 27 Haematinics | "ferritin" or "iron" or Folate or "B12" or "B9" | 1417 |
| 28 | #1 AND #27 | 73 |
| 29 | #2 AND #28 | 51 |
| Additional search | | |
| 30 Potassium blood levels | Potassium or (serum and (potassium or electrolyte*)) or (K and (level* or test*)) or hyperkal?emia or hypokal?emia | 12957 |
| 31 | #1 AND #30 | 493 |
| 32 | #2 AND #31 | 362 |
| 33 Sodium blood levels | Sodium or (serum and (sodium or electrolyte*)) or Na or (Na and (level* or test*)) or hypernatremia or hyponatremia | 7097 |
| 34 | #1 AND #33 | 452 |
| 35 | #2 AND #34 | 369 |

### Table S4. Rapid review 4a: Statins and the side effects

Side effects for statins that can monitored by blood tests including liver disease and liver function tests, Hba1c and diabetes, FBC and anaemia, CK and muscle pain

**Database: KSR Evidence**

| # | Terms | Hits |
|---|---|---|
| 1 | Statin* or "hydroxymethylglutaryl-CoA reductase inhibitors" or "anticholesteremic agents" or simvastatin or rosuvastatin or pravastatin or | 1529 |

| | | |
|---|---|---|
| | atorvastatin or Fluvastatin or “lipid lowering agent*” or “cholesterol lowering agent*” | |
| 2 | (CKD or “kidney function*” or “kidney failure*” or “renal function*” or “renal failure*” or nephropathy or nephropathies or nephrologic* or glomerular filtration rate*” or (kidney AND (disorder* or disease* or dysfunction* or chronic* or insufficienc* or impair*)) or (renal AND (disorder* or disease* or dysfunction* chronic* or insufficienc* or impair*)) or (“kidney failure*” NOT (graft* or allograft* or transplant*))) NOT ((renal NEAR (dialysis or surgery or haemodialysis or hemodialysis or transplant)) OR (kidney NEAR (dialysis or surgery or h?emodialysis or transplant))) | 5481 |
| 3 | #1 AND #2 | 105 |
| 4 | anaem* or anem* or “iron-poor blood” or “Low blood” or “Tired blood” or iron or ferritin or folate or “folic acid” or cobalamin or “total iron” or “iron level*” or “levels of iron” or “iron store*” or “stored iron” or “heme iron” or “haem iron” or ((vitamin or iron or “blood count” or “ferritin or folate or “folic acid” or “cobalamin”) AND (deficien* or low or insufficien* or lack or level*)) | 5097 |
| 5 | #3 AND #4 | 4 |
| 6 | “liver function test*” or “Albumin” or “Alkaline phosphatase” or ALP or “Alanine aminotransferase” or “Alanine transaminase” or ALT or “Aspartate aminotransferase” or “Aspartate transaminase” or AST or Bilirubin or “Gamma glutamyl transpeptidase” or “Gamma-glutamyltransferase” or GGT or “Total protein” or “Serum globulin” or “Liver enzymes” or “Albumin and total protein” or “L-lactate dehydrogenase” or LD or “Prothrombin time” or PT or NAFLD or “liver function” or “fatty liver” or cirrhosis or cirrho* or steatohepatitis or fibrosis or fibro* or “non-alcoholic steatohepatitis” or NASH or hepatic* or hepato* or (hepatic AND (disorders or complications or co-morbid*)) | 11373 |
| 7 | #3 AND #6 | 13 |
| 8 | HbA1c or “h?emoglobin A1” or “glycated h?emoglobin” or “glycated ha?emoglobin” or “glycosylated Hb” or GHb or “plasma glucose” or “total A1” or glycoh?emoglobin or HgbA1c or Hb1c or “blood glucose” or “fasting glucose” | 4058 |
| 9 | #3 AND #8 | 5 |
| 10 | CK or “creatine kinase” or “creatine phosphokinase” or CPK or “CK total” or “CK creatine” or phosphokinase or myalgia or Myopathy (muscle AND (ache* or pain or complaint* or cramp* or weakness)) | 2284 |
| 11 | #3 AND #10 | 10 |
| 12 | “CRP” or “c-reactive protein” or “c reactive protein” | 2349 |
| 13 | #3 AND #12 | 6 |

### Search strategies primary research

#### Table S5. Rapid review 1b: Which test should be used to monitor chronic kidney disease stage 3?

**Databases:**

Embase <1974 to 2023 January 31>
Ovid MEDLINE(R) ALL <1946 to January 31, 2023>

| # | Terms | Hits |
|---|---|---|
| 1 | (CKD or kidney function* or kidney failure* or renal function* or renal failure* or nephropathy or nephropathies or nephrologic or (kidney and (disorder* or disease* or dysfunction* or chronic* or insufficienc* or impair*)) or (renal and (disorder* or disease* or dysfunction* chronic* or insufficienc* or impair*))).ti. | 394591 |

| | | |
|---|---|---|
| 2 | ((kidney or renal) and (graft* or allograft* or transplant*)).ti. | 191883 |
| 3 | ((kidney or renal) and (dialys* or surger* or h?emodialys*)).ti. | 47088 |
| 4 | 2 or 3 | 232731 |
| 5 | 1 not 4 | 349345 |
| 6 | (eGFR or e-GFR or cGFR or c-GFR or (creatinin* adj2 clear*) or glomerul* filtration rate* or glomerulofiltration rate*).mp. | 443724 |
| 7 | ((urea adj4 (blood? or nitrogen*)) or BUN).mp. | 114859 |
| 8 | 5 and 6 and 7 | 4892 |
| 9 | remove duplicates from 8 | 3784 |
| 10 | (monitor* or test* or manag* or screen* or diagnos* or routine or check* or maintain* or control*).mp. | 39295716 |
| 11 | 9 and 10 | 3187 |

#### Table S6. Rapid review 2b and 3b: combined search

- Rapid Review 2b: How prevalent are these acute or chronic complications in chronic kidney disease stage 3 pts compared to general population?
- Rapid review 3b: How prevalent are abnormal test results in chronic kidney disease stage 3 patients compared to general population?

**Database: EMBASE and MEDLINE**

| # | Search component | Terms |
|---|---|---|
| 1 | Chronic kidney disease | (CKD or kidney function* or kidney failure* or renal function* or renal failure* or nephropathy or nephropathies or nephrologic* or glomerular filtration rate* or (kidney and (disorder* or disease* or dysfunction* or chronic* or insufficienc* or impair*)) or (renal and (disorder* or disease* or dysfunction* chronic* or insufficienc* or impair*))).ti. |
| 2 | Primary care/health care databases; UK/European registries | (CRPD or Clinical Practice Research Data* or (GOLD adj3 (data* or regist*)) or AURUM or General Practice Research Data* or GPRD or Health Improvement Network* or THIN data* or QResearch or Q-Research or NHANES or (National Health adj Nutrition Examination Survey) or (Information System? adj3 (Development and Research and Primary Care)) or SIDIAP or Caserta or BIFAP or Base de Datos para la Investigacion Farmacoepidemiologica en Atencion Primaria or ((Finnish or Danish or Swedish) adj3 (national or regional or health*) adj3 (registr* or register)) or DNPR or ((electronic Data Research adj2 Innovation Service) or eDRIS) or (Integrated Primary Care Information Data* or IPCI) or (((Quintiles or IMS) adj Disease Analys*) or mediplus) or ((Quintiles* or IMS or LPD) adj3 health adj3 data*) or ((Quintiles* or IMS) adj3 (longitudinal or patient) adj3 data*) or ((Norwegian or Polish or Czech or MODY) adj3 (Registr* or register)) or Pedianet Data* or (Securite Sociale adj2 Maladie) or (German adj2 (Pharmacoepidemiolog* or Pharmaco-epidemiolog*) adj3 Data*) or Secure Anonymi#ed Information Link* or (ALSPAC or (Avon longitudinal adj3 (cohort or study))) or National Child Development Study or British Cohort Study or (Next Steps adj3 data*)).mp. |
| 3 | Cohorts | ((British or Britain? or United Kingdom? or UK? or Ireland? or Irish or Scotland? or Scottish or Wales? or Welsh or Belgian or Belgium? or Czech* or Danish or Denmark? or Dutch or Netherlands or Europe* or Finnish or Finland? or German* or French or France? or Italian or Italy? or Polish or Poland? or Spain* or Swedish or Sweden? or Swiss or Switzerland?) adj3 (nation* or regional or |

| | | |
|---|---|---|
| | | health* or patient? or population?) adj3 (registr* or register or longitudinal study or cohort study or birth cohort or longitudinal cohort)).mp. |
| 4 | | (((primary care or GP) adj3 database) or (national patient adj (database or regist*))).mp. |
| 5 | | Electronic Health Record*.mp. or EHR.ab. |
| 6 | | 2 or 3 or 4 or 5 |
| 7 | | 1 and 6 |
| 8 | | ((animal model* or mouse or mice or murine* or rat or rats or rodent* or muridae or murids or rabbit* or leporine* or leporidae or guineapig* or cavies or caviidae or hamster* or cricetidae or gerbil* or gerbillinae or cat or cats or feline* or felidae or dog or dogs or canine* or canidae or pig or pigs or piglet* or minipig* or swine* or porcine* or suidae or horse or horses or donkey or donkies or burros or equine* or equidae or sheep or lamb or lambs or ovine or ovidae or goat or goats or cow or cows or cattle or bovine* or bovidae or primate* or monkey or monkeys or macaque or macaques or marmoset or marmosets) not human*).ti. |
| 9 | | (((gestational or maternal* or pregnan*) adj5 hypertensi*) or eclamp* or pre-eclamp* or preeclamp*).ti,kf,hw. |
| 10 | | (((gestational or maternal* or pregnan*) adj3 complication?) and (cardio* or hypertensi*)).ti,kf,hw. |
| 11 | | ((gestational or maternal* or pregnan*) and (HELLP syndrom* or h?emolysis elevated liver enzymes low platelet count*)).ti,kf,hw. |
| 12 | | (covid or covid19 or covid-19 or covid2019 or covid-2019 or ncov* or novel coronavirus or novel betacoronavirus or sars-ncov-2 or sars-cov-2 or postcovid* or longcovid*).ti,kf,hw. |
| 13 | | ((infant* or child* or schoolchild*) not adult*).ti. |
| 14 | | 8 or 9 or 10 or 11 or 12 or 13 |
| 15 | | 7 not 14 |
| 16 | Hba1c | HbA1c or “h?emoglobin A1*” or “glycated h?emoglobin” or “glycated ha?emoglobin” or “glycosylated Hb” or GHb or “plasma glucose” or “total A1*” or glycoh?emoglobin or HgbA1c or Hb1c |
| 17 | diabetes | ((diabet* adj5 (type 2 or type II)) or diabetes mellitus type 2 or diabetes mellitus type II or T2DM or T2 DM or DMT2 or DM T2 or adult onset diabet* or late onset diabet* or matur* onset diabet* or NIDDM or non insulin* diabet* or noninsulin* diabet* or insulin independent diabet* or ketosis resistant diabet*) |
| 18 | | 16 and 17 |
| 19 | | 15 and 18 |
| 20 | | Remove duplicates from 19 |
| 21 | Clotting tests | “Clotting test*” or “Coagulation Test*” or “Prothrombin time” or “Partial thromboplastin time” or “Thrombin time” or INR or “Factor V assay” or “Fibrinogen level” or PT or PT-INR or “Platelet count” |
| 22 | Bleeding disorders | (((bleeding or platelet) and (disorder* or problem*)) or platelet function defects or Disseminated intravascular coagulation or DIC or Prothrombin deficienc* or h?emophilia or Glanzmann disease or Idiopathic thrombocytopenic purpura or ITP or Von Willebrand disease).mp. or (factor and deficiency).ti. |
| 23 | | 21 or 22 |
| 24 | | 15 and 23 |
| 25 | | Remove duplicates from 24 |

| | | |
|---|---|---|
| 26 | Natriuretic Peptide Tests | "Natriuretic Peptide Tests" or BNP or "NT-proBNP" or "brain natriuretic peptide" or "N-terminal pro b-type natriuretic peptide" or "B-type natriuretic peptide" or "brain natriuretic factor" or BNF |
| 27 | Heart failure | "Heart failure" or "cardiac failure" |
| 28 | | 26 or 27 |
| 29 | | 15 and 28 |
| 30 | | Remove duplicates from 29 |
| 31 | Liver disease tests | "liver function test*" or Albumin or "Alkaline phosphatase" or ALP or "Alanine aminotransferase" or "Alanine transaminase" or ALT or "Aspartate aminotransferase" or "Aspartate transaminase" or AST or Bilirubin or "Gamma glutamyl transpeptidase" or "Gamma-glutamyltransferase" or GGT or "Total protein" or "Serum globulin" or "Liver enzymes" or "Albumin and total protein" or "L-lactate dehydrogenase" or LD or "Prothrombin time" or PT |
| 32 | Liver disease | NAFLD or "liver function" or "fatty liver" or cirrhosis or cirrho* or steatohepatitis or fibrosis or fibro* or "non-alcoholic steatohepatitis" or NASH or hepatic* or hepato* or (hepatic AND (complications or co-morbid*)) |
| 33 | | 31 and 32 |
| 34 | | 15 and 33 |
| 35 | | Remove duplicates from 34 |
| 36 | Blood count | FBC or "full blood count" or "complete blood count" or CBC or "Eosinophil count" or Haemoglobin or "Mean corpuscular haemoglobin" or "Mean corpuscular haemoglobin concentration" or "Mean corpuscular volume" or "Monocyte count" or "Neutrophil count" or "All platelet tests" or "Red blood cell count" or "Total white blood cell count" or "Lymphocyte count" or "Packed cell volume" or "Basophil count" or "Differential white cell count" or "Red blood cell size" |
| 37 | Anaemia | (anaem* or anem* or iron-poor blood or Low blood or Tired blood or iron or ferritin or folate or folic acid or vitamin B9 or B12 or cobalamin or total iron or iron level* or levels of iron or iron store* or stored iron or heme iron or haem iron or ((vitamin or iron or ferritin or folate or folic acid or vitamin B9 or B12 or cobalamin) and (deficien* or low or insufficien* or lack or levels))).ti. |
| 38 | | 36 and 37 |
| 39 | | 15 and 38 |
| 40 | | Remove duplicates from 39 |
| 41 | Bone profile | "Bone profile" or "Serum inorganic phosphate" or Calcium or "Bone studies" or "Alkaline phosphatase" |
| 42 | Bone profile diseases | (parathyroid* or hypoparathyroidism or hyperparathyroidism or PTH or parathormone or parathyrin or Pseudohypoparathyroidism or ((bone or bones or cartilage* or chondro* or osteo* or spine or spinal or skeletal or extraskeletal or Ewing) and (cancer* or carcinoma* or neoplas* or metast* or sarcoma* or tumo?r*)) or chondrosarcoma* or vitamin D or insufficien* vitamin D or Cholecalciferol or ergocalciferol).ti. |
| 43 | | 41 and 42 |
| 44 | | 15 and 43 |
| 45 | | Remove duplicates from 44 |
| 46 | Thyroid function tests | Thyroxine or T4 or "Thyroid stimulating hormone" or TSH or "Thyroid function" or "tri-iodothyronine" or T3 |
| 47 | Thyroid disorders | (thyroid disease or thyroid disorders or thyroid dysfunction or dysfunctional thyroid or hypothyroidism or underactive thyroid or hyperthyroidism or hyper-thyroid* or overactive thyroid or Hashimoto* or thyroiditis or Graves* or thyroxin* or thyroid stimulating hormone).mp. |
| 48 | | 46 and 47 |
| 49 | | 15 and 48 |
| 50 | | Remove duplicates from 49 |
| 51 | Lipid tests | "Serum cholesterol" or "total cholesterol" or" High density lipoprotein" or HDL or "Low density lipoprotein" or LDL or Triglyceride* or "HDL:LDL ratio" or |

| | | |
|---|---|---|
| | | “HDL LDL ratio” or “Lipid Profile” or “Fasting Lipid*” or “Non fasting Lipid*” or “Cholesterol Test*” |
| 52 | CVD | (myocardial infarction or stroke* or brain ischemia or cerebrovascular accident or (cardiac adj5 (death or sudden or mortality)) or apoplex* or Abnormal heart rhythm* or arrhythmia* or marfan syndrome or Deep vein thrombosis or pulmonary embolism or heart attack or cardiomyopathy or intracranial h?emorrhage* or coronary syndrome or (cardiovascular adj5 (event* or mortality or death*)) or ((disease* or disorder*) adj5 (cardiovascular or coronary or heart or artery or cardiac or aorta or Congenital heart or Coronary artery or Heart muscle or Heart valve or pericardial or Peripheral vascular or Rheumatic heart or Vascular or blood vessel or isch?emic heart or cerebrovascular))).ti. |
| 53 | | 51 and 52 |
| 54 | | 15 and 53 |
| 55 | | Remove duplicates from 54 |

#### Table S7. Rapid review 4b: Statins and the side effects

Side effects for statins that can monitored by blood tests for FBC and anaemia

**Database: EMBASE and MEDLINE**

| # | Terms |
|---|---|
| 1 Statins | Statin* or "HMG-CoA reductase inhibitor*" or "hydroxymethylglutaryl-CoA reductase inhibitor*" or "anticholesteremic" or simvastatin or rosuvastatin or pravastatin or atorvastatin or Fluvastatin or "lipid lowering" or "cholesterol lowering" |
| 2 Full blood count | FBC or “full blood count” or “complete blood count” or CBC or “Eosinophil count” or Haemoglobin or “Mean corpuscular haemoglobin” or “Mean corpuscular haemoglobin concentration” or “Mean corpuscular volume” or “Monocyte count” or “Neutrophil count” or “All platelet tests” or “Red blood cell count” or “Total white blood cell count” or “Lymphocyte count” or “Packed cell volume” or “Basophil count” or “Differential white cell count” or “Red blood cell size” |
| 3 anaemia | (anaem* or anem* or iron-poor blood or Low blood or Tired blood or iron or ferritin or folate or folic acid or vitamin B9 or B12 or cobalamin or total iron or iron level* or levels of iron or iron store* or stored iron or heme iron or haem iron or ((vitamin or iron or ferritin or folate or folic acid or vitamin B9 or B12 or cobalamin) and (deficien* or low or insufficien* or lack or levels))) |
| 4 | #1 AND #2 AND #3 |
| 5 Clinical trials | ((Clinical trial or RCT) (Clinical Trials.mp OR Clinical Trial Publication Type .mp OR Clinical Trial Data Commons.mp OR Clinical Trials Randomized.mp OR Controlled Clinical Trial Publication Type .mp OR Clinical trial participant person .mp OR Drug Evaluation.mp OR Phase 2 Clinical Trials.mp OR Phase 3 Clinical Trials.mp OR Phase I Clinical Trials.mp OR Clinical Trial Phase II.mp OR Clinical Trial Data Commons 2102D.mp)) Drug Evaluation.mp |
| 6 Adverse effects | (Adverse effects.mp OR "Assessment of adverse drug reactions".mp OR Adverse drug reaction prevention management.mp OR Monitoring Medication Side Effect.mp OR "At risk of medication side effect".mp OR "Adverse reaction to drug".mp OR "Care for Medication Side Effects".mp) "Adverse reaction to drug".mp |
| 7 | #5 AND #6 |
| 8 | #4 AND #7 |

### PRISMA Flow Diagrams

#### Figure S1. Rapid review 1a: Which test should be used to monitor chronic kidney disease?

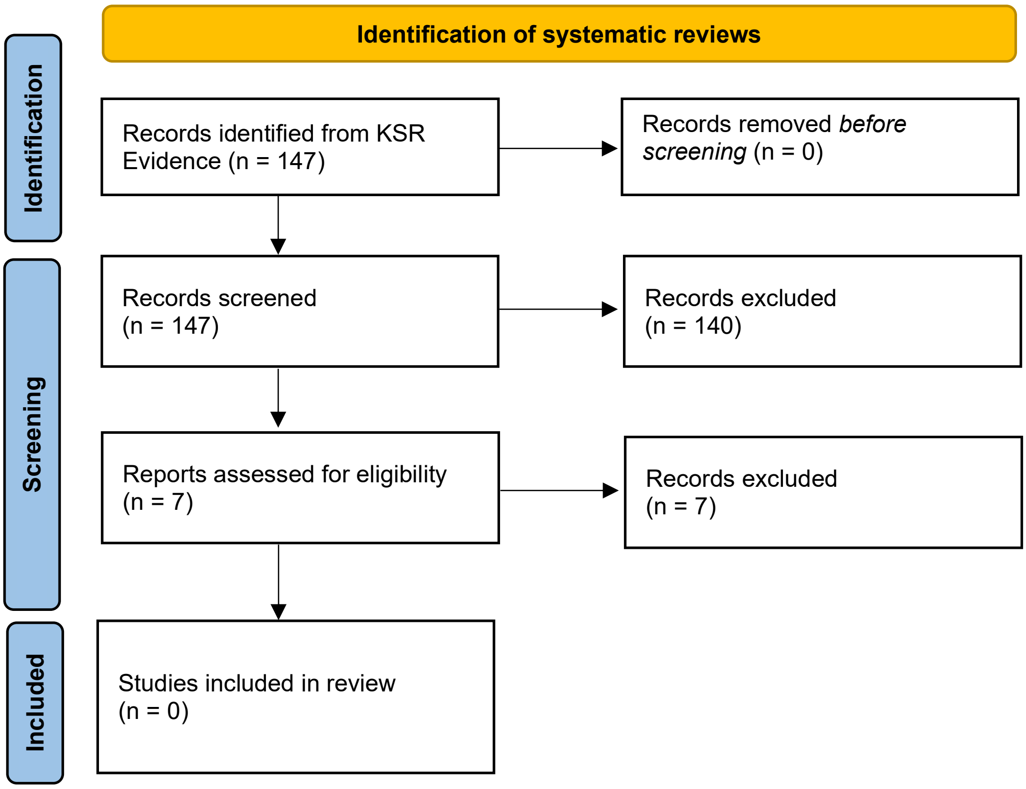

Figure S2. Rapid review 2a: How prevalent are the following acute or chronic complications chronic kidney disease stage 3 patients compared to general population?

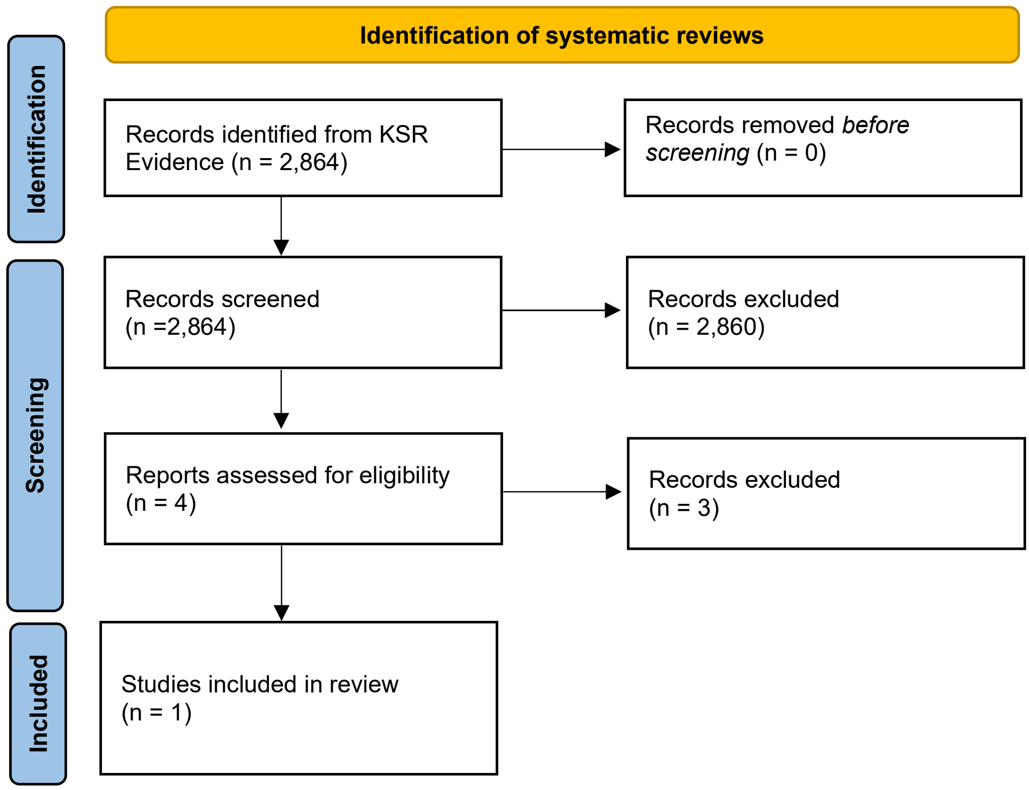

Figure S3. Rapid review 3a: How prevalent are abnormal test results in chronic kidney disease stage 3 patients compared to the general population?

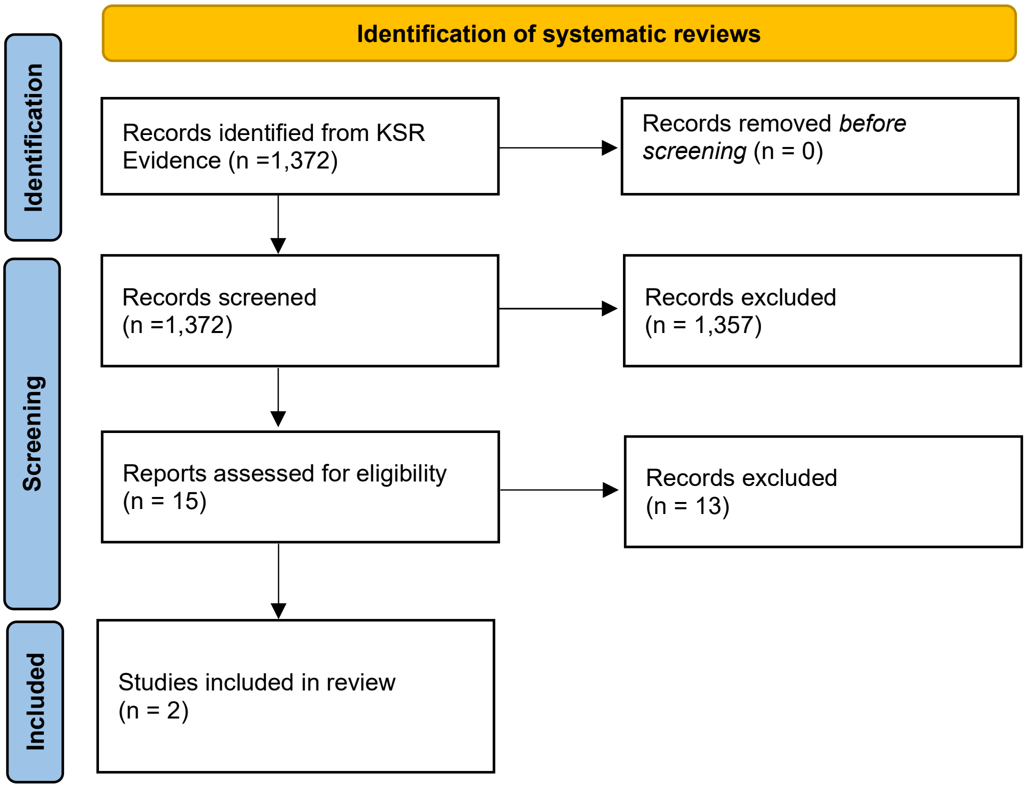

Figure S4. Rapid review 4a: Statins and the side effects

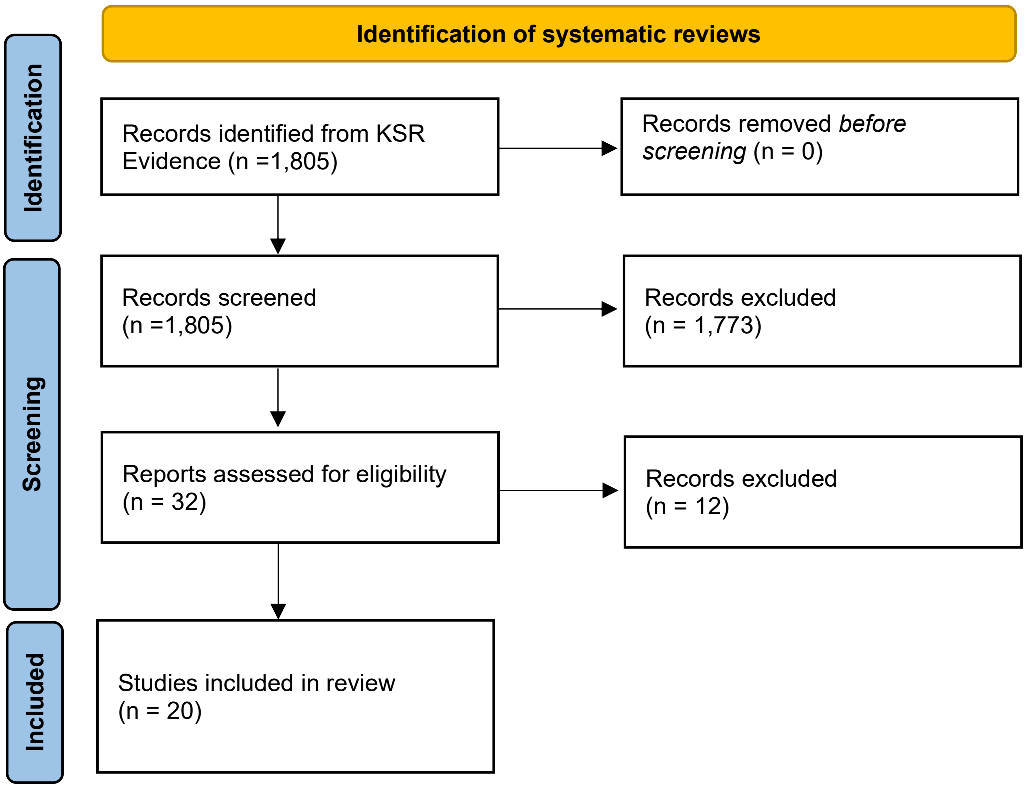

Figure S5. Rapid review 1b: Which test should be used to monitor chronic kidney disease stage 3?

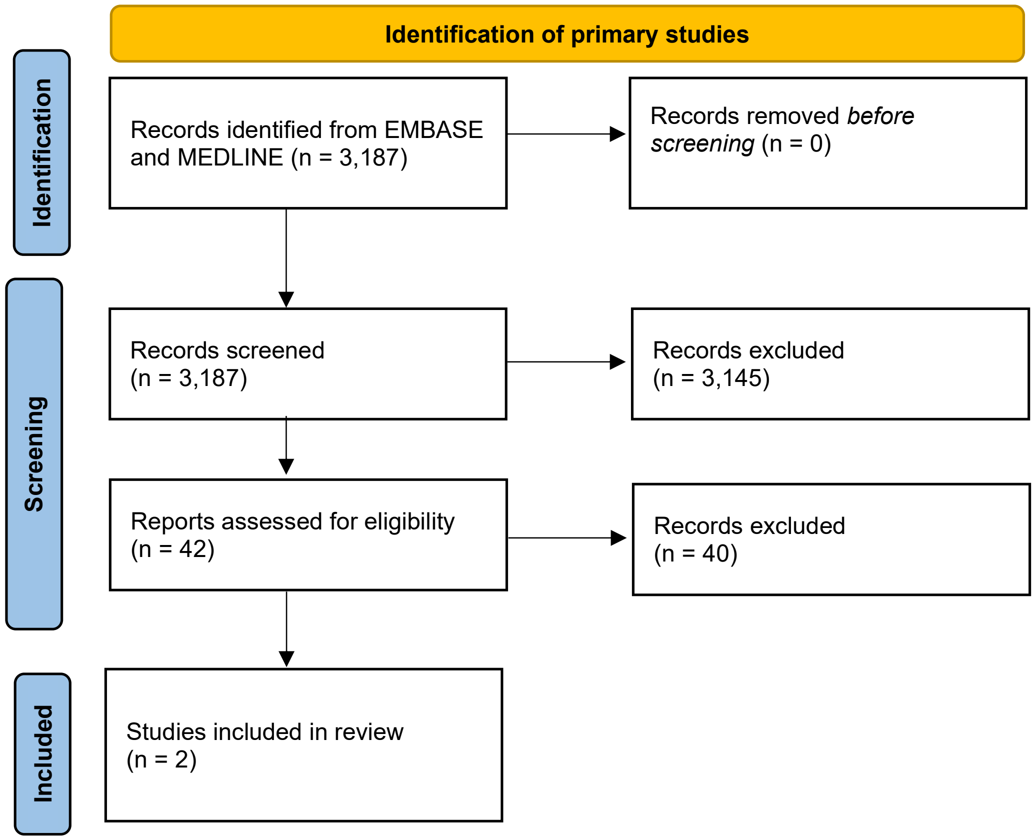

Figure S6. Rapid review 2b and 3b combined: Prevalence of acute or chronic complications and abnormal test results in chronic kidney disease stage 3 patients compared to general population?

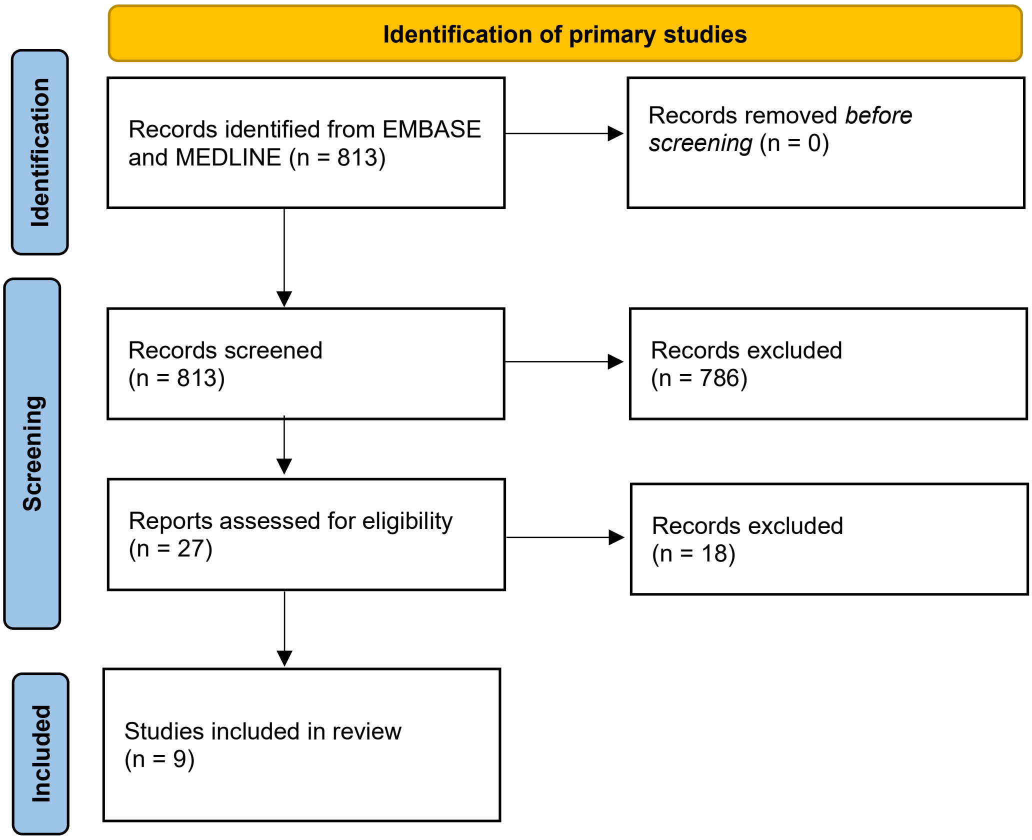

Figure S7. Rapid review 4b: Statins and the side effects

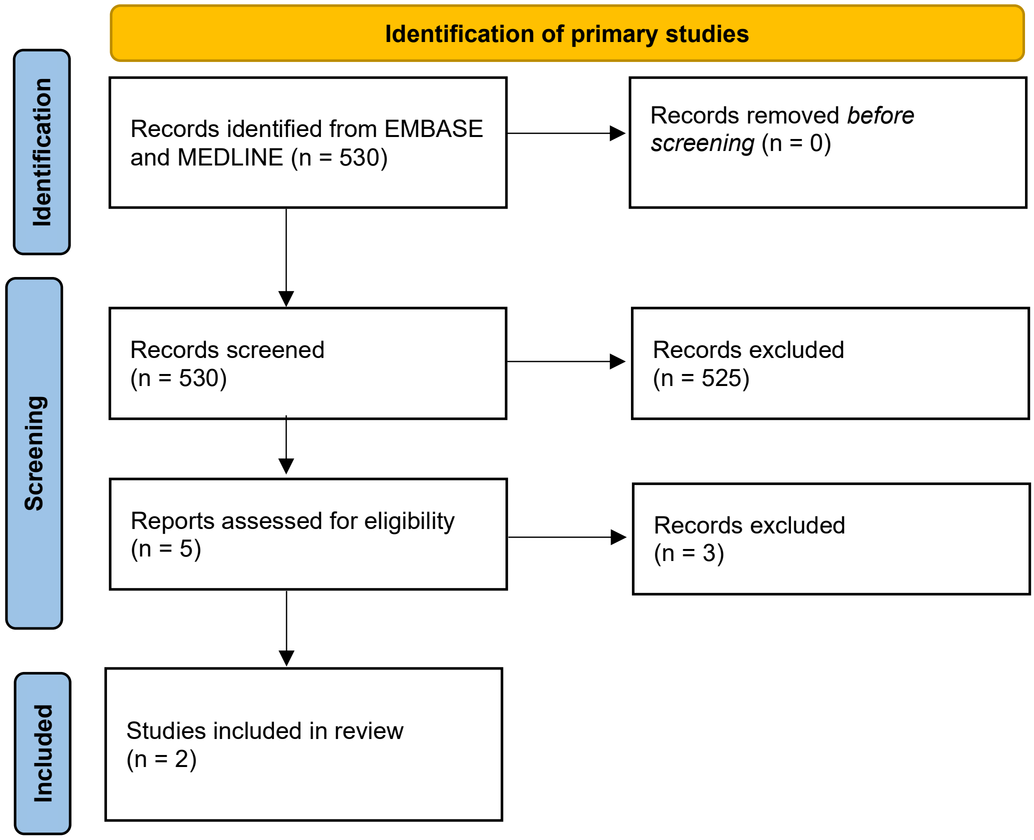

### Evidence report: Chronic Kidney Disease

Authors: Rachel O'Donnell, Catalina Lopez Manzano, Jessica Watson

#### Report Summary

##### Purpose of the consensus meeting

We selected 11 test panels to consider for a routine tests for people with Chronic Kidney Disease (CKD). The purpose of the consensus meeting is to decide which tests should be included for a **minimal testing panel** for people with CKD stage 3 (stage 3a and stage 3b), which tests can be removed from this panel, and for which tests more evidence is needed before a decision can be made.

This minimal testing panel is aimed at an adult that has been diagnosed with a long term condition without complications. This panel lists tests that should be used to regularly monitor this long term condition as standard.

##### Changes made due to the previous consensus feedback

Tests for both C-reactive protein (CRP) and erythrocyte sedimentation rate (ESR) are general markers for inflammation (and infection). It was decided at the previous consensus meeting where T2DM was discussed, that these tests should be removed from future panels as there is no guidance on how to treat low grade inflammation and the tests do not point to a reason for inflammation.

For this report, each testing panel has been split into its constituent individual tests where possible; an overall evidence rating has been given to each individual test and to the testing panel as a whole. We will initially vote on whether to include the "testing panel"; where panels are voted for inclusion or for further evidence, we will then vote on each individual test.

##### Method for Evidence Rapid Reviews

1. Tests were selected based on:
    a. Tests that are currently ordered for patients with chronic kidney disease (CKD) in primary care (CPRD analysis).
    b. Tests that GPs say they would order for the average CKD patient (online survey of GPs).
    c. Tests that are recommended by UK guidelines for routine monitoring of patients with CKD.
2. Tests were categorised as
    a. Tests to monitor disease progression and treatment response
    b. Tests to screen for secondary conditions
    c. Tests to screen for adverse treatment effects
3. We determined a list of questions that needed to be answered with 'yes' for a test to be a useful monitoring test.

a. For Long term condition monitoring:
   I. Can you intervene to slow or prevent further progression of Chronic Kidney Disease?

   II. Can you intervene to improve treatment response in patients with Chronic Kidney Disease?

III. Is there a benefit to earlier intervention?
IV. Which test should be used to monitor renal function?

b. For secondary condition monitoring:
   I. Which acute or chronic complications do these tests pick up?
   II. Is X (or abnormal X tests) more common in LTC patients compared to the general population?
   III. Is there anything the GP can do to manage or treat X?
   IV. Are there clear benefits of earlier detection or treatment?
c. For drug side effect monitoring:
   I. Is X more common in people who take antihypertensive drugs?
   II. Is there anything the GP can do to manage or treat X?
   III. Are there clear benefits of earlier detection or treatment?

4. We looked at the following sources and extracted any evidence in favour or against using each test. We only proceeded to the next source if insufficient evidence was found.
   a. NICE guidelines and NICE clinical knowledge summaries (including references) (CKS)
   b. Systematic reviews
   c. Primary studies in large primary care datasets, cohorts, or relevant trials.

#### Tests for long term condition monitoring summary

There are 2 main blood tests to monitor CKD: blood urea nitrogen (BUN) and estimated globular filtration rate (eGFR). (see Table S8 for summary)

#### Tests to screen for secondary conditions summary

10 testing panels were included to screen for secondary conditions related to CKD. These testing panels were broken down into singular tests within the panel and given and overall evidence rating which will be presented at the consensus meeting for consideration. Evidence was gathered for both prevalence of condition and evidence of using singular tests to test for secondary condition. (see Table S9 for summary)

#### Tests to monitor adverse treatment effects summary

There are no drugs to treat CKD, however there are frequently prescribed drugs to reduce the risk of common secondary conditions. The most frequently prescribed drugs for those with CKD are statins and antihypertensives (angiotensin-converting enzyme (ACE) inhibitors, Angiotensin II receptor blockers (ARB)), which reduce the risk of CVD events; these are included in this report. Antihypertensive drugs ACEs and ARBs were included in the Hypertension report. For this report, we focus on statins as many CKD patients are currently prescribed these, with prescription of statins being actively encouraged for all patients with CKD. We considered the side effects listed in the British National Formulary (BNF) and any with good evidence that appears during the rapid review searches. (see Table S10 for summary)

#### Evidence Rating Guide

We labelled evidence found in the rapid reviews as follows:

| **Rapid review evidence rating** | |
|---|---|
| Good evidence | High quality systematic reviews or high quality studies with large sample sizes |
| Moderate evidence | A systematic review with some quality concerns or several primary studies showing similar results but some studies have quality concerns. |
| Weak evidence | A single primary study with a small sample size with or without quality concerns. |
| Conflicting evidence | Evidence found has contradicting findings. |
| No evidence found | No evidence was found for the condition, adverse effect or single test during the rapid reviews. |
| Unsure | Further discussion is needed in the consensus meeting about the test. |

#### Table S8: Summary Evidence for Drug Adverse Effect Monitoring

| Long term condition | Can you intervene to slow or prevent further progression of Chronic Kidney Disease? | Can you intervene to improve treatment response in patients with Chronic Kidney Disease? | Is there a benefit to earlier intervention? | Which test should be used to monitor renal function? | Evidence rating for single test |
|---|---|---|---|---|---|
| **Chronic Kidney Disease** | Yes | Yes | Yes | eGFR | Good |
| | | | | BUN | Moderate |

#### Table S9: Summary Evidence for tests to screen for secondary conditions

| Blood Test Panel | Related secondary condition | Evidence rating for condition | Individual test types within testing panels | Evidence rating for single test | Treatable/ manageable by GP? | Benefit to earlier detection? |
|---|---|---|---|---|---|---|
| **Blood electrolyte tests** | Hyper- or Hypokalaemia | Weak | Serum Potassium | Weak | Yes | Yes |
| | Hyper- or Hyponatremia | No evidence | Serum Sodium | No evidence | No | Yes |
| **HbA1C** | Type 2 diabetes | No evidence | HbA1C | No evidence | Yes | Yes |
| **Liver Function Tests** | Non-alcoholic fatty liver disease | Weak | Albumin | No evidence | No | No |
| | | | Alkaline phosphatase (ALP) | Weak | No | No |
| | | | Alanine aminotransferase (ALT) | Weak | No | No |
| | | | Aspartate aminotransferase (AST) | No evidence | No | No |
| | | | Bilirubin | No evidence | No | No |
| | | | Gamma glutamyl transpeptidase (GGT) | No evidence | No | No |
| | | | Total protein | No evidence | No | No |
| **Lipid Profile** | Dyslipidaemia, Cardiovascular Disease risk | Weak for Dyslipidaemia | Serum cholesterol | Weak | Yes | Yes |
| | | | High-density lipoprotein (HDL) | Weak | Yes | Yes |
| | | Good for CVD Risk | Low-density lipoprotein (LDL) | Unsure | Yes | Yes |
| | | | HDL:LDL ratio | Unsure | Yes | Yes |
| | | | Triglycerides | No evidence | Yes | Yes |
| **Full blood count** | Anaemia | Good | Haemoglobin (Hb) | Good | Yes | Yes |

| Blood Test Panel | Related secondary condition | Evidence rating for condition | Individual test types within testing panels | Evidence rating for single test | Treatable/ manageable by GP? | Benefit to earlier detection? |
|---|---|---|---|---|---|---|
| **Haematinics** | Anaemia and deficiencies | Weak for serum ferritin | Vitamin B12 | No evidence | Yes | Yes |
| | | | Serum ferritin | Weak | Yes | Yes |
| | | | Folate tests | No evidence | Yes | Yes |
| **Thyroid function tests** | Hyper- or hypothyroidism | Moderate for hypothyroidism | Thyroxine (T4) | Moderate | Yes | Yes |
| | | | Free thyroxine (Free T4) | Moderate | Yes | Yes |
| | | | Thyroid stimulating hormone | No evidence | Yes | Yes |
| | | | Free tri-iodothyronine (T3) | No evidence | Yes | Yes |
| | | | Serum T3 level | No evidence | Yes | Yes |
| **Clotting Tests** | Bleeding disorders | No evidence | Prothrombin time | No evidence | No evidence | No evidence |
| | | | Partial thromboplastin time | No evidence | No evidence | No evidence |
| | | | Thrombin time | No evidence | No evidence | No evidence |
| **Bone Profile** | Late stage kidney disease, bone disorders, serum phosphate levels and parathyroid disorders | Moderate | Serum inorganic phosphate | Moderate | Yes | No evidence |
| | | | Alkaline phosphatase | Moderate | Yes | No evidence |
| | | | Calcium | Weak | Yes | No evidence |
| | | | Calcium adjusted | Weak | Yes | No evidence |
| | | | Parathyroid hormone | Moderate | Yes | No evidence |
| | | | Vitamin D | Weak | Yes | No evidence |
| **B-type natriuretic peptide (BNP)** | Heart failure | Weak | BNP | No evidence | Yes | No evidence |

#### Table S10: Summary Evidence for Drug Adverse Effect Monitoring

| Blood Tests Panel | Adverse Effect | Evidence rating for AE | Individual test types within testing panels | Evidence rating for single test and adverse effect | Treatable/ manageable by GP? | Benefit to earlier detection? |
|---|---|---|---|---|---|---|
| **Blood Glucose tests** | Increased blood glucose or new onset diabetes | Weak (conflicting evidence) | HbA1c | No evidence | Yes | Yes |
| **Renal function tests** | Kidney injury | Weak (conflicting evidence) | eGFR (serum creatine) | No evidence | No | Yes |
| | | | Blood Urea | No evidence | No | Yes |
| | | | Serum potassium | No evidence | No | Yes |
| **Liver function tests** | Liver injury | Good | Albumin | No evidence | No | Yes |
| | | | Alkaline phosphatase (ALP) | No evidence | No | Yes |
| | | | Alanine aminotransferase (ALT) | Good | No | Yes |
| | | | Aspartate aminotransferase (AST) | Good | No | Yes |
| | | | Bilirubin | No evidence | No | Yes |
| | | | Gamma glutamyl transpeptidase (GGT) | No evidence | No | Yes |
| | | | Total protein | No evidence | No | Yes |
| **Anaemia** | Anaemia | Weak evidence against AE | Haemoglobin (Hb) | Weak | Yes | Yes |
| | | | Vitamin B12 | Weak | Yes | Yes |
| | | | Serum ferritin | Weak | Yes | Yes |
| | | | Folate tests | Weak | Yes | Yes |
| **Creatine Kinase** | Muscle symptoms, myalgia, muscle pain, or rhabdomyolysis | Good | Creatine Kinase | Weak (conflicting evidence) | No | Yes |

#### Tests to monitor disease progression and treatment response

##### eGFR (serum creatine), and urea blood tests (BUN)

*Can you intervene to slow or prevent further progression of Chronic Kidney Disease?*

Yes, monitoring CKD, assessing for underlying causes or conditions that may worsen the disease, such as hypertension, prescribing or altering drug prescriptions and optimizing lifestyle can slow progression of CKD.

| **Source** | **Evidence** | **Quality Concerns** |
|---|---|---|
| CKS NICE | "People with a declining estimated glomerular filtration rate (eGFR) and progressive CKD have a worse prognosis, increased risk of complications, and require early intervention to prevent further renal deterioration compared with people with a stable eGFR. There is evidence that delayed referral leads to increased morbidity and mortality in people with advanced CKD [KDIGO, 2013].<br>A reduction in blood pressure in people with CKD has the potential dual benefits of reducing risk of disease progression and reducing cardiovascular risk [NICE, 2015a; Fraser, 2016]. Expert opinion in a review article notes that prevention of CKD disease progression includes management of hypertension, appropriate use of renin-angiotensin system antagonists, and avoidance of nephrotoxic drugs where possible [Vassalotti, 2016].<br>https://cks.nice.org.uk/topics/chronic-kidney-disease/management/management-of-chronic-kidney-disease/ | |
| Shabaka 2021 [1] | "Therapeutic Insights in CKD Progression delay CKD progression or even reverse it. In addition to RAS blockade, SGLT2 inhibitors and bicarbonate therapy have proved to retard CKD progression, and new drugs targeting fibrosis and inflammation, as well as regenerative therapy may enhance these effects in our goal to mitigate CKD progression and consequently cardiovascular and global mortality." | No |
| Molina 2021 [2] | "Close monitoring to adherence to dietary recommendations and frequent evaluation of nutritional status is fundamental in the management of patients with CKD, since it can affect important health outcomes, including CKD progression, quality of life, morbidity, and mortality. Within these nutritional measures, salt restriction, Low Protein Diet and with Amino Acids and Keto Analogs, have been shown in recent meta-analyses of RCTs to be effective in modifying the natural history of CKD, delaying the fall of the GFR, decreasing proteinuria, BP levels, or bone mineral disorder parameters, without increasing the risk of Protein Energy Wasting." | no |

*Can you intervene to improve treatment response in patients with Chronic Kidney Disease?*

Yes, there are no medications to treat CKD, but there are many medications to treat the associated conditions that help slow progression of CKD related secondary conditions.

| Source | Evidence |
| --- | --- |
| CKS NICE | The NICE guideline development group noted that the beneficial effects of angiotensin-converting enzyme (ACE) inhibitors and angiotensin-II receptor antagonists (AIIRAs) appeared to be more closely related to the presence or absence of proteinuria rather than blood pressure control.<br>This approach is supported by expert opinion in a review article which notes that greater early reductions in proteinuria are associated with slower progression of kidney disease [Webster, 2017].<br><br>For all people with CKD, prescribe lipid-lowering therapy with a statin, for the primary or secondary prevention of CVD. Be aware that renal impairment is a risk factor for myopathy and rhabdomyolysis adverse effects of statins. For people who have an eGFR of 30 mL/min/1.73 m2 or less, consider seeking specialist advice before increasing statin doses.<br><br>https://cks.nice.org.uk/topics/chronic-kidney-disease/management/management-of-chronic-kidney-disease/ |

*Is there a benefit to earlier intervention?*

Yes, detecting CKD earlier can prevent and slow progression of secondary conditions and CKD.

| Source | Evidence |
| --- | --- |
| CKS NICE | A reduction in blood pressure in people with CKD has the potential dual benefits of reducing risk of disease progression and reducing cardiovascular risk [NICE, 2015a; Fraser, 2016]. Expert opinion in a review article notes that prevention of CKD disease progression includes management of hypertension, appropriate use of renin-angiotensin system antagonists, and avoidance of nephrotoxic drugs where possible [Vassalotti, 2016].<br><br>https://cks.nice.org.uk/topics/chronic-kidney-disease/management/management-of-chronic-kidney-disease/ |
| Shlipak 2020 [3](KDIGO) | “The KDIGO Controversies Conference participants were unanimous that the bulk of evidence supports systematic approaches to screen for, risk stratify, and treat persons with CKD. Because interventions to slow CKD progression and reduce cardiovascular risk are evidence based and have been shown to improve outcomes, conference attendees agreed that the focus should be on strategies to maximize deployment of CKD screening, risk stratification, and treatment efforts.” |

*Which test should be used to monitor renal function?*

Estimated globular filtration rate (eGFR) is the standard test to measure renal function. There are multiple equations to calculate eGFR from serum creatine, however, for the UK, CKD-EPI is recommended to use. However, some studies show that BUN is independently associated with renal decline and can be used to inform renal outcomes.

| **Source** | **Tests** | **Evidence** | **Quality Concerns** |
|---|---|---|---|
| NICE guidelines | eGFR | Monitor renal function by checking serum creatinine and estimated glomerular filtration rate (eGFR) together with urinary albumin: creatinine ratio (ACR) to identify 'accelerated progression' of CKD. https://cks.nice.org.uk/topics/chronic-kidney-disease/management/management-of-chronic-kidney-disease/ | |
| NICE guideline NG203 | eGFR equations | NICE recommends the use the Chronic Kidney Disease Epidemiology Collaboration (CKD-EPI) creatinine equation to estimate GFR creatinine for adults. Evidence for this recommendation can be found: https://www.nice.org.uk/guidance/ng203/evidence/a-diagnostic-accuracy-of-egfr-calculations-in-adults-children-and-young-people-from-black-asianand-other-minority-ethnic-groups-with-ckd-pdf-9204445982 | |
| Systematic reviews | | | |
| None identified | | | |
| Primary studies | | | |
| Lin 2019 | blood urea nitrogen to creatinine ratio (BCR) vs eGFR | Population: Taiwanese population<br>Sample size: 2,685 (CKD n=801)<br>Findings:<br>- High blood urea nitrogen to creatinine ratio (BCR) indicates various physiological conditions (including age, dehydration drug use)<br>- A BCR of 20 or greater caused misestimation of the CKD stage diagnosis (by eGFR equations MDRD and CKD-EPI). GFR estimates for patients with high BCR should be interpreted cautiously<br>**Conclusion: BUN may be used to inform CKD stage, but not monitor CKD itself.** | Only Taiwanese population included. BUN was used as BCR. |
| Seki 2019 | BUN and eGFR | Population: Japanese population with CKD stage 3-5<br>Sample size: 459 (stage 3 (n= 161), stage 4 n= (178), stage 5 (n = 120)<br>Findings:<br>- higher BUN levels were associated with adverse renal outcomes independent of eGFR in patients with moderate to severe CKD.<br>- there was no significant interaction for renal outcomes between BUN and eGFR levels. These results might suggest that the impact of the BUN level on kidney disease progression is stronger at earlier stages of CKD.<br>**Conclusion: BUN levels may be associated with renal outcomes, independent of kidney function.** | Only Japanese population included. |

### Tests to screen for secondary conditions

#### **Blood Electrolyte tests** (Serum potassium and serum sodium)

*Which acute or chronic complications do these tests pick up?*

Hyper/Hypokalaemia and Hyper/Hyponatremia (CKS).

*Are abnormal blood electrolyte levels more common in CKD patients compared to the general population?*

Evidence suggests the prevalence of hyperkalaemia/high serum potassium levels increases as renal function declines. No evidence was found for the prevalence of high serum sodium levels in CKD patients vs the general population.

| **Source** | **Blood Tests** | **Evidence** | **Quality Concerns** |
|---|---|---|---|
| NICE Guidelines | Electrolytes (potassium and sodium) | Persistent hyperkalaemia is a suspected complication of CKD. Specialist management of and specialist dietary advice about potassium can be offered.<br>https://cks.nice.org.uk/topics/chronic-kidney-disease/management/management-of-chronic-kidney-disease/<br>Important to monitor serum potassium levels before starting renin–angiotensin system antagonist.<br>https://www.nice.org.uk/guidance/ng203/resources/chronic-kidney-disease-assessment-and-management-pdf-66143713055173 | |
| *Systematic reviews* | | | |
| None identified | | | |
| *Primary studies* | | | |
| Bhan 2010 [4] | Serum potassium | • Population: US population between 2002 to 2004<br>• Sample size: 69,215 (non-CKD: 60,743; CKD: 8,472, (75% stage 3a, 21% stage 3b, and 4% stage 4)<br>• Findings:<br>○ Potassium levels (mg/dL) CKD: 4.2 (± 0.3) vs non-CKD: 4.0 (± 0.3) P = < 0.0001<br>• **Conclusion: mean potassium levels increase slightly in CKD patients compared to those without CKD (at stage 3).** | Results were not divided into CKD stages (However, 96% of the CKD participants had stage 3, so results are assumed for stage 3 CKD) |
| Hsu 2002 [5] | Serum Potassium | • Population: NHANES 1988-1994, patients aged 17+.<br>• Sample size: 14,722 (women n = 7,835, men n = 6,887).<br>• Findings:<br>• Changes in Potassium (mmol/L)<br>○ CrCl > 70 ml/min: Women: 0.0 [95% CI 0.0–0.0]; Men: 0.0 [95% CI 0.0–0.1]<br>○ CrCl > 60 ml/min: Women: 0.0 [95% CI 0.0–0.1]; Men: 0.0 [95% CI 0.0–0.1]<br>○ CrCl > 50 ml/min: Women: 0.1 [95% CI 0.0–0.1]; Men: 0.0 [95% CI 0.0–0.2] | Study uses Creatinine clearance (CrCL) Stages rather than eGFR stages |

| Source | Blood Tests | Evidence | Quality Concerns |
|---|---|---|---|
| | | ○ CrCl > 40 ml/min: Women: 0.1 [95% CI 0.0–0.1]; Men: 0.0 [95% CI 0.0–0.2]<br>○ CrCl > 30 ml/min: Women: 0.1 [95% CI 0.1–0.2]; Men: 0.0 [95% CI 0.1–0.03]<br>○ CrCl > 20 ml/min: Women: 0.2 [95% CI 0.1–0.3]; Men: 0.0 [95% CI 0.2–0.05]<br>○ CrCl < 20 ml/min: Women: 0.4 [95% CI 0.1–0.7]; Men: 0.0 [95% CI 0.0–0.5]<br>**Conclusion: there is a stepwise increase in mean serum potassium level as renal function decreases. This is more apparent in women than men.** | |

*Is there anything the GP can do to manage or treat abnormal blood electrolyte levels?*

Yes, mild or moderate hyperkalaemia may be managed by dietary modification or medication changes for those with hypertension. However, in the event of life-threatening acute hyperkalaemia, emergency hospital admission may be needed, and drugs are available to reduce potassium levels to a safer range.

| Source | Evidence |
|---|---|
| NICE | Sodium zirconium cyclosilicate for treating hyperkalaemia<br>NICE Technology appraisal guidance [TA599]<br>Published: 24 January 2022<br>https://www.nice.org.uk/guidance/ta599/resources/sodium-zirconium-cyclosilicate-for-treating-hyperkalaemia-pdf-82607272135621<br><br>Patiromer for treating hyperkalaemia<br>NICE Technology appraisal guidance [TA623]<br>Published: 13 February 2020<br>https://www.nice.org.uk/guidance/ta623/resources/patiromer-for-treating-hyperkalaemia-pdf-82609015577029<br>Both State:<br>**Treatment for hyperkalaemia depends on its severity. Life-threatening acute hyperkalaemia needs emergency treatment in hospital.**<br>Small rises in serum potassium above this can cause electrocardiogram (ECG) changes. To lower the risk of cardiac arrest, clinicians use active potassium-lowering treatments, then identify and remove the cause of hyperkalaemia. The guidelines include the following treatments:<br>• calcium chloride or calcium gluconate intravenously to protect the heart if there is ECG evidence of hyperkalaemia<br>• insulin and glucose intravenously to move potassium from the blood into cells<br>• nebulized salbutamol as an adjunctive therapy to insulin and glucose for serum potassium levels of 6.5 mmol/litre and above to move potassium from the blood into cells<br>• after severe hyperkalaemia has resolved, potassium-binding agents for 3 or more days (namely, calcium resonium given orally) to remove potassium from the body<br>• stopping or reducing RAAS inhibitors, which can increase serum potassium levels.<br><br>**The aim of treatment for chronic hyperkalaemia is to lower potassium levels to prevent acute life-threatening hyperkalaemia. Treatment includes:** |

| | |
|---|---|
| | • advising people with chronic kidney disease to avoid foods high in potassium<br>• stopping or reducing RAAS inhibitors and potassium-sparing diuretics<br>• avoiding non-steroidal anti-inflammatory drugs and trimethoprim. |

*Are there clear benefits of earlier detection or treatment?*

Yes. Late detection of high potassium levels can lead to increased risk of cardiac arrest.

#### HbA1C

*Which acute or chronic complications do these tests pick up?*

Type 2 diabetes mellitus (T2DM) (NICE guidance)

*Is T2DM (or abnormal blood glucose tests) more common in CKD patients compared to the general population?*

No evidence for type 2 diabetes as a secondary condition for CKD. Nice guidelines recommends that during initial assessment for CKD, the individual is tested for diabetes.

| **Source** | **Blood Tests** | **Evidence** | **Quality Concerns** |
|---|---|---|---|
| NICE Guidelines | There are multiple possible causes and risk factors for chronic kidney disease (CKD) and its progression, including: Diabetes mellitus. https://cks.nice.org.uk/topics/chronic-kidney-disease/background-information/causes/<br>If a diagnosis of chronic kidney disease (CKD) is suspected, arrange initial investigations in primary care. Check the person's nutritional status, body mass index (BMI), blood pressure, and serum HbA1c and lipid profile to assess for cardiovascular risk. https://cks.nice.org.uk/topics/chronic-kidney-disease/diagnosis/initial-investigations/#classification-of-ckd | | |
| *Systematic reviews* | | | |
| None identified | | | |
| *Primary Studies* | | | |
| None identified | | | |

*Is there anything the GP can do to manage or treat T2DM?*

Yes, Lifestyle changes and drug treatment to control blood glucose levels. There are several options for medications which can be changed in dose or combination.

*Are there clear benefits of earlier detection or treatment?*

Yes, early intervention with drugs has long term benefits through lowering blood glucose and reducing the complications of Diabetes.

**Liver function tests** (albumin, alkaline phosphatase (ALP), alanine aminotransferase (ALT), aspartate aminotransferase (AST), bilirubin, gamma glutamyl transpeptidase (GGT), total protein)

*Which acute or chronic complications do these tests pick up?*

Non-alcoholic fatty liver disease (NAFLD), including non-alcoholic fatty liver (NAFL) and non-alcoholic steatohepatitis (NASH, NICE).

*Is NAFLD (or abnormal liver test results) more common in CKD patients compared to the general population?*

No, one study suggests that NAFLD, and raised ALP and ALT levels are not significantly different to the general population.

| **Source** | **Blood Tests** | **Evidence** | **Quality Concerns** |
|---|---|---|---|
| NICE Guidelines | Not Mentioned | | |
| *Systematic Reviews* | | | |
| None identified | | | |
| *Primary Studies* | | | |
| Deng 2021 [6] | ALP, alkaline phosphatase; ALT, alanine aminotransferase | • Population: NHANES 2017-2018<br>• Sample size: 4,869 (Non-CKD (n = 3,151), CKD (n = 815)<br>• Findings:<br>○ Fibrosis-4 (FIB-4) score: non-CKD: 1.0 (0.70) vs CKD 1.6 (1.0), OR: 1.23 [95% CI 1.05 - 1.45; p = 0.011].<br>○ NAFLD fibrosis score: non-CKD: −1.7 (1.4) vs CKD −0.40 (1.6), OR: 1.09 [95% CI 0.98-1.20; p = 0.102].<br>○ ALT IU/L: non-CKD 22.2 (15.9) vs CKD 20.8 (17.0), OR: 1.00 [95% CI 1.00-1.00; p = 0.231].<br>○ ALP IU/L: non-CKD 78.2 (27.0) VS CKD 85.8 (30.2), OR: 1.00 [95% CI 1.00-1.01; p = 0.010].<br>**Conclusion: there is no significant difference between CKD patients and non-CKD individuals for prevalence of NAFLD, fibrosis, or raised ALT and ALP levels.** | Not all sampled patients were included, and exclusion could be associated with outcome. |

*Is there anything the GP can do to manage or treat NAFLD?*

No. Other than lifestyle advice (which is same recommends for patients with CKD) there is not much the GP can do. This suggests there is no benefit to monitor CKD patients for NAFLD.

<table>
<tr><th>Source</th><th>Evidence</th></tr>
<tr><td>NICE</td><td>Non-alcoholic fatty liver disease (NAFLD): assessment and management<br>NICE guideline [NG49]<br>Published: 06 July 2016<br>https://www.nice.org.uk/guidance/ng49/evidence/full-guideline-pdf-2548213310<br><br>• Weight reduction interventions: no relevant studies identified. Nevertheless, NICE recommends offering advice on physical activity and diet to people with NAFLD.<br>• Dietary modification and supplements: Do not offer omega-3 fatty acids to adults with NAFLD because there is not enough evidence to recommend their use.<br>• Exercise interventions: There is some evidence that exercise reduces liver fat content.<br>• Lifestyle modification: there is some evidence that lifestyle interventions (diet, behavioural modifications, and exercise) are beneficial (although the effect may be small). NICE recommends considering lifestyle interventions regardless of the patients' BMI.<br>• Alcohol advice: Recommendation to alcohol consumption within national limits. More research is needed to determine whether reducing alcohol intake below national limits is beneficial for NAFLD patients.<br>• No evidence to make any recommendations on caffeine and fructose intake.<br>Pharmacological interventions: there is currently no licensed treatment for NAFLD.</td></tr>
</table>

**Lipid profile** (Serum cholesterol, high-density lipoprotein (HDL), low-density lipoprotein (LDL), triglycerides, HDL:LDL ratio)

*Which acute or chronic complications do these tests pick up?*

Dyslipidaemia (abnormal levels of different types of lipids in the blood) is a risk factor for CVD (NICE), including stroke and myocardial infarction, and ischemic heart disease. Lipid tests are used to calculate the risk of CVD (QRISK 3 [7]), which use total cholesterol and HDL measurements.

*Are dyslipidaemia and cardiovascular risk more common in CKD patients compared to the general population?*

One study suggests total cholesterol levels increase in CKD patients as the disease progresses, with stage 3 CKD patients having the highest total cholesterol levels. The same study also reported that HDL levels have a non-significant decline at stage 4 of CKD. CKD patients have a higher risk of CVD events than non-CKD individuals, with risk increasing further with declining renal function.
Total cholesterol and HDL levels are used to calculate CVD risk, therefore there is good evidence to include them. However, unsure about LDL and HDL:LDL ratio as no evidence was found for those tests, but they can be calculated alongside total cholesterol and HDL levels.

| **Source** | **Blood Tests** | **Evidence** | **Quality Concerns** |
|---|---|---|---|
| NICE Guidelines | People with CKD are 5–10 times more likely to die prematurely of cardiovascular disease than they are to progress to end-stage renal disease (ESRD). Economic modelling estimated that in England in 2009–10, an extra 7,000 strokes and 12,000 myocardial infarctions occurred in people with CKD, compared with age- and gender-matched controls [NICE, 2015a]. The risk of death rises exponentially as renal function worsens, and is largely secondary to cardiovascular disease [Webster, 2017]. https://cks.nice.org.uk/topics/chronic-kidney-disease/background-information/complications/ | | |
| *Systematic reviews* | | | |
| Yuan 2022 [8] | No specific blood tests mentioned | - Population: Chinese population without history of stroke<br>- Sample size: 89 studies (40 prospective cohorts, n = 1,984,552) that reported effect estimates of the risk factors of first stroke occurrence in Chinese population<br>- Findings:<br> o CKD was associated with increased risk of stroke (RR = 1.65 [95%CI 1.36–2.01; $p < 0.001$]).<br>**Conclusion: the risk of stroke is significantly higher in those with CKD.** | Only Chinese population included, insufficient search, and no definition for CKD. |
| Masson 2015 [9] | No specific blood tests mentioned | - Population: Global and European General population<br>- Sample size: 83 studies; 63 cohort studies (2,085,225 participants) and 20 RCTs (168,516 participants)<br>- Findings<br> o GFR of <90mL/min/1.73m$^2$ was associated with an increased risk of all-cause stroke by 39% (RR: 1.39 [95% CI 1.31–1.47]). | No |

| Source | Blood Tests | Evidence | Quality Concerns |
|---|---|---|---|
| | | ○ In participants with a GFR of 60–90 mL/min/1.73 $m^2$ the risk of stroke was increased by 10% (RR: 1.10 [95% CI 1.03–1.19]), by 43% in participants with a GFR of 30–60 mL/min/1.73 $m^2$ (RR: 1.43 [95% CI 1.33–1.54]) and by 70% in participants with an GFR of <30 mL/min/1.73 $m^2$ (RR: 1.70 [95% CI 1.47–1.96; test for difference; p < 0.001]).<br>○ For every 10 mL/min/1.73 $m^2$ decrease in GFR (relative to the reference group >90 mL/min/1.73 $m^2$), the risk of having a stroke increased by 7% (RR: 1.07 [95% CI 1.04–1.09]).<br>**Conclusion: The risk of all-cause stroke increased further with declining renal function. With the increase being 10% for stage 2, 43% for stage 3a and 3b, and 70% in stage 4 and up.** | |
| *Primary Studies* | | | |
| Borg 2022 conference abstract [10] | No specific blood test mentioned. | • Population: Danish health registries based in Copenhagen<br>• Sample size: 171,133 individuals included (CKD 1 or 2 (control group) n = 157,002)).<br>• Findings:<br>○ In general, event rates were low in CKD stages 1 and 2 and rose with higher stages of CKD.<br>○ MI HR: Stage 3 CKD 1.3 [95% CI 1.22 – 1.4], stage 4 CKD 2.15 [95% CI 1.73 – 2.68], stage 5 2.4 [95% CI 1.2-4.81].<br>○ Stroke HR: Stage 3 CKD 1.11 [95% CI 1.06 – 1.16], stage 4 CKD 1.2 [95% CI 1.01 – 1.44], stage 5 1.67 [95% CI 0.97- 2.89].<br>○ CVD death HR: Stage 3 CKD 1.26 [95% CI 1.21 – 1.31], stage 4 CKD 2.05 [95% CI 1.81 – 2.31], stage 5 2.67 [95% CI 1.8 -3.96].<br>**Conclusion: There is an increased risk of CVD events, stroke, MI and CVD death in those with CKD, which rises with stage. CVD deaths and MI have a more significant increase than risk of stroke in CKD patients.** | Population from Copenhagen, small area of representation. Hospitalisation used for outcomes. |
| Inker 2012 [11] | Total cholesterol and HDL cholesterol. | • Population: NHANES 1988-1994 and 1999-2006<br>• Sample size: 30,528<br>• Findings:<br>○ Total cholesterol (mg/dL): No CKD: 201.9 (±0.6), stage 1: 207.7 (±1.8), stage 2: 213.4 (±2.0), stage 3: 217.6 (±2.1), stage 4: 207.9 (± 6.6), p = < 0.001.<br>○ HDL (mg/dL): No CKD: 51.7 (±0.2), stage 1: 51.2 (±0.7), stage 2: 51.8 (±0.7), stage 3: 51.1 (± 0.4), stage 4: 48.9 (±1.7), p = 0.2.<br>**Conclusion: total cholesterol levels significantly increase as stages of CKD increase, peaking at stage 3, then lowering at stage 4. HDL levels slightly decline at stage 4, but no significant decrease is seen between stages.** | Not known if medication was adjusted for. |

*Is there anything the GP can do to manage or treat this?*

Yes, the GP can advise on diet and lifestyle changes,  tight blood pressure control through lifestyle change and antihypertensive drugs, and offering lipid modification therapy (usually statins). However, as CVD events are the most common cause of death in the CKD patient population, NICE recommend that statins are offered to all patients with CKD to reduce the risk of CVD events, irrespective of the results of lipid blood tests.

| Source | Evidence |
|---|---|
| NICE | **For all people with CKD, prescribe lipid-lowering therapy with a statin, for the primary or secondary prevention of CVD.**<br>People with CKD are at increased risk of CVD and will benefit from CVD risk modification including the use of lipid-modifying therapy. The NICE guideline development group noted the trial evidence on statin use in CVD has largely excluded people with CKD. It found no evidence to assume that the effectiveness of lipid-modifying therapy would be different for people with CKD compared with the general population [NICE, 2015a].<br>https://cks.nice.org.uk/topics/chronic-kidney-disease/management/management-of-chronic-kidney-disease/<br><br>**Lipid modification - CVD prevention: Scenario: Lipid therapy - primary prevention of CVD**<br>Last revised in May 2021<br>https://cks.nice.org.uk/topics/lipid-modification-cvd-prevention/management/lipid-therapy-primary-prevention-of-cvd/<br>Consider offering lipid modification therapy (without the need for a formal risk assessment) to people who aged 85 years or over (particularly people who smoke or have raised blood pressure) |

*Are there clear benefits of earlier detection or treatment?*

Yes. Untreated elevated lipid levels increase the risk of CVD.

| Source | Evidence |
|---|---|
| NICE | **Lipid modification - CVD prevention:**<br>**What is the relationship between blood lipids and cardiovascular health?**<br>Last revised in May 2021<br>https://cks.nice.org.uk/topics/lipid-modification-cvd-prevention/background-information/lipids-cardiovascular-health/<br>“Total cholesterol is an important predictor of CVD events. However, non-high density lipoprotein cholesterol (non-HDL-C), the difference between total and HDL cholesterol, which usually makes up 60–70% of total serum cholesterol is a powerful risk factor for CVD.”<br>“Elevated triglyceride levels (greater than 2.3 mmol/L), especially when HDL-C levels are low, is a risk factor for CVD and is independent of total cholesterol.” “Extreme levels of triglycerides (greater than 20 mmol/L) are associated with pancreatitis and a high risk of morbidity and mortality.” |

#### Full blood count (haemoglobin)

*Which acute or chronic complications do these tests pick up?*

Anaemia (low red blood cell count) (CKD).

*Is anaemia more common in CKD patients compared to the general population?*

Yes, as low haemoglobin levels are more common in CKD patients. At stage 3 anaemia significantly more prevalent compared to non-CKD and CKD stage 1 and 2. At stage 4 there is a large increase in prevalence of anaemia compared to stage 3.

| Source | Blood Tests | Evidence | Quality Concerns |
|---|---|---|---|
| NICE Guidelines | Renal anaemia (haemoglobin less than 11 g/dL [110 g/L]) [NICE, 2015a]. This may present with symptoms such as tiredness, shortness of breath, lethargy, and palpitations. It may be due to reduced production of erythropoietin by the kidney, reduced red blood cell survival, and iron deficiency [Webster, 2017].<br>https://cks.nice.org.uk/topics/chronic-kidney-disease/background-information/complications/<br>In adults: if eGFR is above (G2) 60 ml/min/1.73 m2, investigate other causes of anaemia as it is unlikely to be caused by CKD. If eGFR is between (G3A+B) 30 and 60 ml/min/1.73 m2, investigate other causes of anaemia, but use clinical judgement to decide how extensive this investigation should be, because the anaemia may be caused by CKD. If eGFR is below 30 ml/min/1.73 m2, think about other causes of anaemia but note that anaemia is often caused by CKD. [2021] (G4+)<br>https://www.nice.org.uk/guidance/ng203/resources/chronic-kidney-disease-assessment-and-management-pdf-66143713055173 | | |
| *Systematic Reviews* | | | |
| None identified | | | |
| *Primary Studies* | | | |
| Stauffer 2014 [12] | Haemoglobin levels | • Population: NHANES 2007-2008 and 2009-2010.<br>• Sample size: 12,077 adult individuals (CKD n = 2,125, non-CKD n = 9,952)<br>• Findings:<br> ○ The prevalence of anaemia was 15.4% [95% CI13.1-18.2] in people with CKD and 6.3% [95% CI 5.3-7.4] in those without.<br> ○ The prevalence of anaemia increased with stage of CKD. Stage 1: 8.4% [95% CI 5.5-12.4], stage 2: 12.2% [95% CI 9.2-12.4], stage 3: 17.4% [95% CI 13.7-21.8], stage 4: 50.3% [95% CI 37.2-63.4], stage 5: 53.4% [95% CI 34.1-71.7].<br>**Conclusion: The prevalence of anaemia increases with CKD progression, with the most significant increase at stage 4.** | No |
| Inker 2012 [11] | Haemoglobin levels | • Population: NHANES 1988-1994 and 1999-2006.<br>• Sample size: 30,528<br>• Findings: | Not known if medication was adjusted for. |

| Source | Blood Tests | Evidence | Quality Concerns |
|---|---|---|---|
| | | ○ The prevalence of anaemia was highest in patients with stages 3 or 4 CKD, and was lower in patients with stage 2 compared to stage 1.<br>○ Risk ratios and 95% CI of related conditions by CKD stage vs non-CKD: Anaemia: stage 1: 1.34 [95% CI 1.04 – 1.74], stage 2: 1.02 [95% CI 0.82 – 1.26], stage 3 2.06 [95% CI 1.79 – 2.37], stage 4: 5.23 p95% CI 3.99 – 6.86], p < 0.001.<br>**Conclusion: There is a significant increase in risk of anaemia in CKD patients at stage 3, which then doubles at stage 4.** | |
| Bhan 2010 [4] | Haemoglobin levels | • Population: US population between 2002 to 2004<br>• Sample size: 69,215 (non-CKD: 60,743; CKD: 8,472, (75% stage 3a, 21% stage 3b, and 4% stage 4)<br>• Findings:<br>○ Haemoglobin (g/dL) CKD: 12.9 (±1.6) vs non CKD: 13.8 (±1.5), p = <0.0001<br>• **Conclusion: Haemoglobin levels decrease in CKD patients compared to those without CKD (at stage 3).** | Results were not divided into CKD stages (However, 96% of the CKD participants had stage 3, so results are assumed for stage 3 CKD) |
| Clase 2007 [13] | Haemoglobin levels | • Population: NHANES 1986 – 1994<br>• Sample size: 15,802<br>• Findings:<br>○ Prevalence of haemoglobin <110g/L: stage 1: 1.5% [95% CI 1.2-1.7], stage 2: 0.9% [95% CI 0.5-1.2], stage 3: 3.5% [95% CI 2.4-4.7], stage 4 and 5: 42.2% [95% CI 28.3-56.0].<br>○ ORs: stage 4 and 5: 58.09 [95% CI 22.41-150.59], stage 3: 2.11 [95% CI 1.24-3.58], stage 2: 0.56 [95% CI 0.34 - 0.95], stage 1 – reference.<br>**Conclusion: There is a significant decrease in haemoglobin levels in stage 3 CKD, and a significant and severe decrease in patients with stage 4 and 5 CKD.** | No |
| Astor 2002 [14] | Haemoglobin levels | • Population: NHANES 1988-1994<br>• Sample size: 15,419<br>• Findings:<br>○ The median haemoglobin level among men decreased from 14.9g/dL at stage 3a to 13.8 g/dL at stage 3b and to 12.0 g/dL at stage 4.<br>○ The median haemoglobin level among women similarly decreased from 13.5 g/dL at an estimated GFR of 60 mL/min per 1.73 $m^2$ to 12.2 g/dL at an estimated GFR of 30 ml/min per 1.73 $m^2$ and to 103 g/dL at an estimated GFR of 15 ml/min per 1.73$m^2$.<br>○ Above an estimated GFR of 90 mL/min per 1.73$m^2$, the median haemoglobin level was mildly but significantly ($p < 0.001$) lower at higher estimated GFRs.<br>**Conclusion: Below 60 mL/min per 1.73 $m^2$ (Stage 3), a lower eGFR was associated with a lower haemoglobin level for men and women.** | No |

*Is there anything the GP can do to manage or treat this?*

Yes, give dietary advice and for iron deficiency anaemia prescribe iron tablets, for Vit B12 deficiency anaemia refer or administer hydroxocobalamin, for folate deficiency anaemia prescribe folic acid.

| Source | Evidence |
|---|---|
| NICE | **Anaemia - iron deficiency: Management (CKS)**<br>Last revised in November 2021<br>https://cks.nice.org.uk/topics/anaemia-iron-deficiency/management/<br>Iron deficiency anaemia:<br>• Prescribe all people with iron deficiency anaemia one tablet once daily of oral ferrous sulfate, ferrous fumarate or ferrous gluconate — continue treatment for 3 months after iron deficiency is corrected to allow stores to be replenished.<br>• Monitor the person to ensure that there is an adequate response to iron treatment.<br><br>**Anaemia - B12 and folate deficiency (CKS)**<br>Last revised in July 2020<br>https://cks.nice.org.uk/topics/anaemia-b12-folate-deficiency/<br>Vit B12 deficiency anaemia:<br>• For people with neurological involvement: Seek urgent specialist advice from a haematologist.<br>• For people with no neurological involvement: Initially administer hydroxocobalamin 1 mg intramuscularly three times a week for 2 weeks. The maintenance dose depends on whether the deficiency is diet related or not. Give dietary advice about foods that are a good source of vitamin B12.<br>Folate deficiency anaemia:<br>• Prescribe oral folic acid 5 mg daily — in most people, treatment will be required for 4 months.<br>• Check vitamin B12 levels in all people before starting folic acid.<br>• Give dietary advice about foods that are a good source of folic acid. |

*Are there clear benefits of earlier detection or treatment?*

Yes, untreated anaemia can worsen long term patient outcomes (however, more evidence or expert input is needed). For example, iron deficiency in patients with low ejection fraction is associated with decreased long-term survival and a lower quality of life.

| Source | Evidence | Quality Concerns |
|---|---|---|
| Wienbergen 2018 [15] | • Population: Germany and Switzerland patients with heart failure with reduced ejection fraction<br>• Sample size: 949 patients (Iron deficiency (n = 505) without iron deficiency (n = 418)<br>• Findings: | No |

| | |
|---|---|
| | ○ Patients with iron deficiency had a higher long-term mortality compared to those without iron deficiency (19.5% vs. 13.7%, p = 0.02) and reported a lower quality of life. Only a minority of patients with ID (9.3%) received iron supplementation during long-term course, just 4.7% intravenously.<br>○ In the adjusted analysis a significant interaction remained, with iron deficiency being a significant predictor of 1-year mortality in patients without anaemia (HR 2.15, 95% CI 1.12–3.78), but not in anaemic patients (HR 0.99, 95% CI 0.65–1.49).<br>• **Conclusion: Anaemia was associated with an elevated mortality.** |

#### **Haematinics** (Vit B12, Serum ferritin, Folate tests)

*Which acute or chronic complications do these tests pick up?*

Vitamin B12, ferritin, and folate deficiencies, also called iron deficiency anaemia, vitamin B12 and folate anaemia.

*Is anaemia, or vitamin B12, ferritin, and folate deficiencies more common in CKD patients compared to the general population?*

One study reported on the risk of low serum ferritin amongst CKD patients. No evidence was found for Vitamin B12 or folate deficiencies.

| **Source** | **Blood Tests** | **Evidence** | **Quality Concerns** |
|---|---|---|---|
| NICE Guidelines | Renal anaemia (haemoglobin less than 11 g/dL [110 g/L]) [NICE, 2015a]. This may present with symptoms such as tiredness, shortness of breath, lethargy, and palpitations. It may be due to reduced production of erythropoietin by the kidney, reduced red blood cell survival, and iron deficiency [Webster, 2017].<br>https://cks.nice.org.uk/topics/chronic-kidney-disease/background-information/complications/ | | |
| *Systematic Reviews* | | | |
| None identified | | | |
| *Primary Studies* | | | |
| Hsu 2002 [5] | Serum ferritin | • Population: NHANES III 1988 – 1994<br>• Sample size: 15,971<br>• Findings:<br>○ 47% of women and 44% of men had serum ferritin <100 ng/ml.<br>▪ CrCl > 80 ml/min: Women: 81 [95% CI 79–84]; Men: 31 [95% CI 29–33]<br>▪ CrCl > 70 ml/min: Women: 81 [95% CI 77–85]; Men: 28 [95% CI 23–34]<br>▪ CrCl > 60 ml/min: Women: 78 [95% CI 74–81]; Men: 36 [95% CI 29–43]<br>▪ CrCl > 50 ml/min: Women: 62 [95% CI 58–67]; Men: 34 [95% CI 27–41]<br>▪ CrCl > 40 ml/min: Women: 55 [95% CI 51–59]; Men: 41 [95% CI 33–50]<br>▪ CrCl > 30 ml/min: Women: 59 [95% CI 53–65]; Men: 37 [95% CI 30–44]<br>▪ CrCl > 20 ml/min: Women: 47 [95% CI 41–54]; Men: 44 [95% CI 33–55] | Study uses creatinine clearance (CrCL) Stages rather than eGFR stages. |

| | | |
|---|---|---|
| | | ▪ CrCl < 20 ml/min: Women: 47 [95% CI 28–67]; Men: 44 [95% CI 28–59]<br>**Conclusion: Frequency of low ferritin (<100 ng/ml) increases as CKD progresses (as defined by CrCL).** |

*Is there anything the GP can do to manage or treat this?*

Yes, give dietary advice and for iron deficiency anaemia prescribe iron tablets, for Vit B12 deficiency anaemia refer or administer hydroxocobalamin, for folate deficiency anaemia prescribe folic acid.

*Are there clear benefits of earlier detection or treatment?*

Yes, untreated anaemia can worsen long term patient outcomes (however, more evidence or expert input is needed). For example, iron deficiency in patients with low ejection fraction is associated with decreased long-term survival and a lower quality of life.

**Thyroid function tests** (Thyroxine, thyroid stimulating hormone, free triiodothyronine (T3), free thyroxine, serum T3 level)

*Which acute or chronic complications do these tests pick up?*

Hyper- or hypothyroidism (CKS)

*Are hyper- or hypothyroidism more common in CKD patients compared to the general population?*

CKD is associated with a higher prevalence of hypothyroidism, both overt and subclinical. One study suggests that prevalence of hyperthyroidism was insignificant in CKD patients.

| **Source** | **Blood Tests** | **Evidence** | **Quality Concerns** |
|---|---|---|---|
| NICE Guidelines | Malignancy. People with ESRD (end-stage renal disease) may have an excess cancer risk, particularly affecting the renal tract and thyroid gland. The exact cause is not known, but risk factors may include exposure to immunosuppressive agents, and immune dysregulation caused by chronic uraemia [Webster, 2017]. | | |
| *Systematic Reviews* | | | |
| None identified | | | |
| *Primary Studies* | | | |
| Siva 2016 [16] | TSH and Free T4 levels | • Population: Indian population<br>• Sample size: 100 (50 healthy and 50 with CKD longer than 6 months pre-dialysis)<br>• Findings:<br>○ Among the CKD patients, subclinical hypothyroidism, hypothyroidism, subclinical hyperthyroidism and hyperthyroidism have their prevalence rates as 40%, 20%, 4% and 2% respectively.<br>○ Subclinical hypothyroidism is more prevalent in CKD patients (40%) when compared with the general population (6%). Subclinical hypothyroidism was found to be 8% stage II, 10% stage III, 20% stage IV and 12% in stage V of CKD patients.<br>○ Prevalence of thyroid disorders are 71.4%, 62.5%, 56% and 80% in the stages II, III, IV and V of CKD respectively.<br>**Conclusion: Prevalence of subclinical hypothyroidism is high among all the stages of CKD patients except for stage 5. Incidence of thyroid abnormalities increases as the stage of CKD advances and eGFR declines.** | Small sample size and no information on enrolment process. Population also from India, so concerns on applicability of data. |
| Lo 2005 [17] | TSH and Free T4 levels | • Population: NHANES 1988- 1994<br>• Sample size: 14,623<br>• Findings: | No |

| Source | Blood Tests | Evidence | Quality Concerns |
|---|---|---|---|
| | | ○ We observed that the prevalence of hypothyroidism was increased in persons with reduced GFR, ranging from 5.4% for persons with estimated GFR ≥90 mL/ min/1.73m$^2$ to more than 20% in persons with estimated GFR <60 mL/min/1.73m$^2$.<br>○ After adjusting for age, sex, and race/ethnicity, compared with estimated GFR≥60 mL/min/1.73m$^2$, GFR<60 mL/min/1.73m$^2$ was associated with an increased odds of hypothyroidism (adjusted odds ratio (OR) 1.58 [95% CI 1.16-2.14]).<br>○ Compared with GFR ≥90 mL/min/1.73m$^2$ (healthy), the adjusted odds of hypothyroidism increased among subjects with lower estimated GFR: adjusted OR 1.07 [95% CI 0.86-1.32] for GFR 60 to 89 mL/min/1.73m$^2$ (stage 2), 1.57 [95% CI 1.11-2.22] for GFR 45 to 59 mL/min/1.73m2 (stage 3a), 1.81 [ 95% CI 1.04-3.16] for GFR 30 to 44 mL/min/1.73m$^2$ (stage 3b), and 1.97 [95% CI 0.69-5.61] for GFR <30 mL/min/1.73m2 (stage 4) (P = 0.008 for trend).<br><br>**Conclusion: The odds of hypothyroidism increase with a decline in eGFR, with the most significant increase seen in patients with stage 3a CKD.** | |

*Is there anything the GP can do to manage or treat hyper- or hypothyroidism?*

Yes, the GP can prescribe levothyroxine for primary or subclinical hypothyroidism, and radioactive iodine for adults with Graves' disease or hyperthyroidism secondary to multiple nodules.

| Source | Evidence |
|---|---|
| NICE | Thyroid disease: assessment and management<br>NICE guideline [NG145]<br>Published: 20 November 2019<br>https://www.nice.org.uk/guidance/ng145<br><br>- Offer levothyroxine as first-line treatment for adults, children and young people with primary hypothyroidism.<br>- Consider levothyroxine for adults with subclinical hypothyroidism who have a TSH of 10 mlU/litre or higher on 2 separate occasions 3 months apart. Follow the recommendations in section 1.4 on follow-up and monitoring of hypothyroidism.<br>- Offer radioactive iodine as first-line definitive treatment for adults with Graves' disease, unless antithyroid drugs are likely to achieve remission (see recommendation 1.6.11), or it is unsuitable (for example, there are concerns about compression, malignancy is suspected, they are pregnant or trying to become pregnant or father a child within the next 4 to 6 months, or they have active thyroid eye disease). |

| | |
|---|---|
| | - Offer radioactive iodine as first-line definitive treatment for adults with hyperthyroidism secondary to multiple nodules unless it is unsuitable (for example, there are concerns about compression, thyroid malignancy is suspected, they are pregnant or trying to become pregnant or father a child within the next 4 to 6 months, or they have active thyroid eye disease). |

*Are there clear benefits of earlier detection or treatment?*

(more evidence or expert input needed)

#### **Clotting tests** (Prothrombin time, Partial thromboplastin time, Thrombin time)

*Which acute or chronic complications do these tests pick up?*

Bleeding disorders (CKS)

*Are bleeding disorders or abnormal clotting test results more common in CKD patients compared to the general population?*

No evidence was identified on the risk of bleeding disorders amongst CKD patients.

| Source | Blood Tests | Evidence | Quality Concerns |
|---|---|---|---|
| NICE Guidelines | Not mentioned | | |
| *Systematic Reviews* | | | |
| None identified | | | |
| *Primary Studies* | | | |
| None identified | | | |

*Is there anything the GP can do to manage or treat this?*

High mean platelet volume or high platelet distribution width do not necessarily require treatment, but they may indicate other problems (more evidence or expert input needed).

| Source | Evidence |
|---|---|
| NICE | Platelets - abnormal counts and cancer:<br>Scenario: Management of platelet count outside the normal range (CKS)<br>Last revised in June 2021<br>https://cks.nice.org.uk/topics/platelets-abnormal-counts-cancer/management/management/<br>- The GP needs to establish the cause underlying the bleeding problems in order to treat them. |

*Are there clear benefits of earlier detection or treatment?*

(more evidence or expert input needed)

#### **Bone profile** (Serum inorganic phosphate, Calcium, Calcium adjusted, Vitamin D, alkaline phosphatase, parathyroid hormone)

*Which acute or chronic complications do these tests pick up?*

Serum calcium and phosphorus levels, vitamin D deficiency, parathyroid problems. (CKS)

*Are bone cancer, vitamin D deficiency, or parathyroid problems (or abnormal bone profile results) more common in CKD patients compared to the general population?*

Evidence suggests a higher risk of hyperphosphatemia and hyperparathyroidism in patients with stage 3 CKD, with a significant increase seen in patients with stage 4 CKD. Serum calcium levels only begin to increase at stage 4 CKD. When stage 3 is broken down into stage 3a and 3b, hyperphosphatemia and hyperparathyroidism are both increased in stage 3b, but no significant increase in serum phosphorus is seen in stage 3a. Similarly, Vitamin D levels are only significantly decreased in stage 3b.

| **Source** | **Blood Tests** | **Evidence** | **Quality Concerns** |
|---|---|---|---|
| NICE Guidelines | Renal mineral and bone disorder. This may present with bone pain, increased bone fragility, or extra-skeletal calcification, such as in the skin or blood vessels [Bello, 2017; KDIGO, 2017; Webster, 2017].<br>It is caused by disturbed vitamin D, calcium, parathyroid hormone (PTH), and phosphate metabolism due to impaired regulation of intestinal absorption and renal tubular excretion. This subsequently causes abnormalities in bone turnover and mineralisation. Serum calcium may be low or high initially, with vitamin D deficiency, raised serum phosphate, low serum calcium, and secondary or tertiary hyperparathyroidism seen in progressive CKD [Vassalotti, 2016; Bello, 2017; Webster, 2017].<br>https://cks.nice.org.uk/topics/chronic-kidney-disease/background-information/complications/<br>Arrange serum calcium, phosphate, vitamin D, and parathyroid hormone tests to exclude renal metabolic and bone disorder for people with CKD category stages 4 or 5. Consider checking serum parathyroid hormone and vitamin D level in people with CKD stages 1, 2, 3a, and 3b if bone disease is suspected clinically, for example, if there is unexplained hypo- or hypercalcaemia. | | |
| *Systematic Reviews* | | | |
| None Identified | | | |
| *Primary Studies* | | | |
| Inker 2012 [11] | Serum PTH Levels and serum phosphate levels | • Population: NHANES 1988-1994 AND 1999-2006<br>• Sample size: 30528<br>• Findings:<br>o There was a progressive increase in the prevalence of hyperparathyroidism with the Stages. The prevalence of hyperphosphatemia was lower at Stage 2 compared to Stage 1 but highest at Stages 3 and 4.<br>o Risk ratios and 95% CI of related conditions by CKD stage vs No CKD: | Not known if medication was adjusted for. |

| Source | Blood Tests | Evidence | Quality Concerns |
|---|---|---|---|
| | | ○ Hyperphosphatemia: stage 1: 0.91 (95% CI [0.70 – 1.18]), stage 2: 1.06 (95% CI [0.75 – 1.50]), stage 3: 1.61 (95% CI [1.36 – 1.92]), stage 4: 3.64 (95% CI [2.40 – 5.52]), p < 0.001.<br>○ hyperparathyroidism: stage 1: 1.15 (95% CI [0.70 – 1.58]), stage 2: 1.00 (95% CI [0.63 – 1.59]), stage 3: 2.63 (95% CI [1.97 – 3.51]), stage 4: 6.05 (95% CI [4.46 – 8.191]).<br><br>**Conclusion: Risk of hyperphosphatemia increases at stage 3 CKD, and triple at stage 4. The risk of hyperparathyroidism doubles at stage 3 and severely increase in stage 4.** | |
| Bhan 2010 [4] | serum phosphate levels and serum calcium levels | • Population: US population between 2002 to 2004<br>• Sample size: 69215 (non-CKD: 60743; CKD: 8472, (75% stage 3a, 21% stage 3b, and 4% stage 4)<br>• Findings:<br>○ Calcium (mg/dl) CKD: 9.3 (±0.4) vs non CKD: 9.3 (±0.4) P=0.47<br>○ Phosphorus (mg/dl) CKD: 3.2 (±0.5) vs non CKD: 3.1 (±0.5) P = <0.0001<br>**Conclusion: there is no difference in calcium levels for CKD patients compared to those without CKD at (stage 3). Phosphorus levels slightly increase in CKD patients compared to those without CKD (at stage 3).** | Results were not divided into CKD stages (However, 96% of the CKD participants had stage 3, so results are assumed for stage 3 CKD) |
| Muntner 2009 [18] | Serum PTH Levels and serum phosphate levels | • Population: NHANES 2003-2004<br>• Sample size: 3,949<br>• Findings: >= 60 ml/min/1.73 m2 (stage 2 or 1)= ref<br>○ Parathyroid hormone (Ipth) (>70 pg/ml)<br>▪ Stage 3a (45 to 59 ml/min/1.73m$^2$) = 2.46 (95% CI [1.77- 3.43])<br>▪ Stage 3b (30 to 44 ml/min/1.73m$^{2)}$ = 5.25 (95% CI [3.13-8.80])<br>○ Serum calcium (<9.4 mg/dl)<br>▪ Stage 3a = 0.95 (95% CI [0.66- 1.37])<br>▪ Stage 3b = 1.18 (95% CI [0.73-1.90])<br>○ Serum phosphorus (>4.2 mg/dl)<br>▪ Stage 3a = 1.34 (95% CI [0.96- 1.85])<br>▪ Stage 3b = 1.54 (95% CI [1.05-2.24])<br>○ Vitamin D (25(OH) D) (<17.6 ng/ml)<br>▪ Stage 3a = 1.26 (95% CI [0.96- 1.65])<br>▪ Stage 3b = 1.58 (95% CI [1.05-2.37])<br>**Conclusion: There is no significant difference in serum calcium levels in CKD stages 1 and 2 vs 3a and 3b. There is a slight increase in serum phosphorus levels at stage 3b. Parathyroid hormone levels significantly increase at both stage 3a and stage 3b, with stage 3b also being significantly higher than stage 3a. Vitamin D levels significantly decline in stage 3b, but not in stage 3a.** | Excluded patients with an eGFR of <30 ml/min/1.73m2 |

| Source | Blood Tests | Evidence | Quality Concerns |
|---|---|---|---|
| Clase 2007 [13] | Serum Calcium and serum phosphate | • Population: NHANES 1986 – 1994<br>• Sample size: 15802<br>• Findings:<br> ○ Prevalence of calcium <2.15 mmol/l: Stage 1: 4.1% (95% CI [3.3-4.8]), Stage 2: 4.0% (95% CI [3.2-4.8]), Stage 3: 3.4% (95% CI [1.7-5.2]) stage 4: 8.2% (95% CI [1.6-14.8])<br> ▪ ORs: stage 4 and 5: 1.57 (95% CI [0.56 -4.41]), stage 3: 0.60 (95% CI [0.35- 1.03]), stage 2: 0.85 (95% CI [0.70- 1.04]), stage 1 – reference<br> ○ Prevalence of phosphate >1.6 mmol/l: Stage 1: 4.3% (95% CI [3.4-5.1]), Stage 2: 3.1% (95% CI [2.0-4.2]), stage 3: 5.7% (95% CI [3.3-8.2]) Stage 4: 32.7% (95% CI [19.6-45.9])<br> ▪ ORs: stage 4 and 5: 150.36 (95% CI [35.19- 642.50]), stage 3: 2.24 (95% CI [0.33 -15.11]), stage 2: 0.37 (95% CI [0.07- 2.02]), stage 1 – reference<br>• **Conclusion: there is a significant increase in phosphorus levels in patients with stage 4 and 5 CKD, and a similar significant decrease in Calcium levels.** | No |

*Is there anything the GP can do to manage or treat this*

Yes, treatments are available for Vitamin D deficiency, phosphate levels, and hyperparathyroidism (and calcium levels through Hyperparathyroidism management).

| Source | Evidence |
|---|---|
| NICE | **Vitamin D deficiency in adults:**<br>**Scenario: Management of vitamin D deficiency or insufficiency (CKS)**<br>Last revised in September 2021<br>https://cks.nice.org.uk/topics/vitamin-d-deficiency-in-adults/management/management/<br>• Advise that oral vitamin D3 is the vitamin D preparation of choice for the treatment of vitamin D deficiency, and vitamin D2 is an alternative option in some clinical situations.<br><br>**Hyperparathyroidism (primary): diagnosis, assessment and initial management**<br>NICE guideline [NG132]<br>Published: 23 May 2019<br>https://www.nice.org.uk/guidance/ng132<br>• Surgical management<br>• Non-surgical management if surgery is unsuccessful. |

| | |
|---|---|
| | **Hyperphosphataemia in chronic kidney disease**<br>**NICE clinical guideline 157, part of managing chronic kidney disease (CG182) and managing anaemia in CKD (NG8)**<br>Published: March 2013<br>https://www.nice.org.uk/guidance/ng203/evidence/full-guideline-2013-hyperphosphataemia-in-ckd-pdf-9206080240<br>• Adults with hyperphosphataemia in chronic kidney disease should be offered a calcium-based phosphate binder as a first-line treatment in addition to dietary management |

*Are there clear benefits of earlier detection or treatment?*

(more evidence or expert input needed)

#### **B-type natriuretic peptide** (BNP)

*Which acute or chronic complications do these tests pick up?*

Heart failure (NICE, CKS)

*Is heart failure more common in CKD patients compared to the general population?*

Studies found that CKD increases the risk of severe heart failure, with one study suggesting a significant increase in stage 3a and a 2 fold increase in stage 3b.

| **Source** | **Blood Tests** | **Evidence** | **Quality Concerns** |
|---|---|---|---|
| NICE Guidelines | Cardiovascular disease such as ischaemic heart disease, peripheral arterial disease, heart failure, and stroke disease [KDIGO, 2013]. People with CKD are 5–10 times more likely to die prematurely than they are to progress to end-stage renal disease (ESRD). The risk of death rises exponentially as renal function worsens, and is largely secondary to cardiovascular disease [Webster, 2017]. https://cks.nice.org.uk/topics/chronic-kidney-disease/background-information/complications/ | | |
| *Systematic Reviews* | | | |
| None Identified | | | |
| *Primary Studies* | | | |
| Borg 2022 conference abstract [10] | No specific blood test mentioned. | • Population: Danish health registries based in Copenhagen<br>• Sample size: 171,133<br>• Findings:<br>○ In general, event rates were low in CKD stages 1 and 2 and rose with higher stages of CKD.<br>○ Heart failure HR: Stage 3 CKD 1.35 (95% CI [1.29 – 1.41]), stage 4 CKD 2.13 (95% CI [1.87 – 2.43]), stage 5 2.23 (95% CI [1.51 -3.57])<br>**Conclusion: there is an increased in risk of heart failure in those with CKD, which rises with stage.** | Population was taken from Copenhagen, small area of representation. Hospitalisation used for outcomes |
| Iwagami 2017 [19] | No specific blood test mentioned. | • Population: UK adult population CPRD<br>• Sample size: 242, 349 matched pairs (N= 242,349 with CKD, N= 242,349 without CKD) (CKD stage 3a (N=172,555), CKD stage 3b (N=55,500), CKD stage 4 or 5 (N=14,294)).<br>• Findings:<br>○ The largest incidence rate difference was seen for heart failure with 6.6/1000 person-years (PY) (due to the incidence rate of 9.7 vs. 3.1/1000 PY in patients with and without CKD)<br>○ Relative risk for heart failure with adjusted hazard ratio of 1.66 (95% CI, 1.59 - 1.75)<br>○ adjusted hazard ratio for heart failure separated by stage: CKD stage 3a: 1.43 (95% CI [1.37 to 1.50]), CKD stage 3b: 2.13 (95% CI [2.02 to 2.24]), CKD stage 4 or 5: 3.29 (95% CI [3.06 to 3.54]).<br>**Conclusion: Patients with CKD have a higher risk of heart failure than those without CKD, increasing with stage.** | Heart failure was reported as hospitalisation for Heart Failure, so only severe cases of HF were included, this may reduce heart failure risk. |

*Is there anything the GP can do to manage or treat this?*

Yes, there are treatments available for heart failure.

| Source | Evidence |
|---|---|
| NICE | **Heart failure - chronic:**<br>**Scenario: Confirmed heart failure with reduced ejection fraction (CKS)**<br>Last revised in August 2021<br>https://cks.nice.org.uk/topics/heart-failure-chronic/management/confirmed-heart-failure-with-reduced-ejection-fraction/<br>For a person with confirmed heart failure and reduced ejection fraction:<br>- Ensure drugs which may cause or worsen heart failure are reviewed and stopped if appropriate.<br>- To relieve symptoms of fluid overload, ensure the person has been prescribed a loop diuretic.<br>- Prescribe both an angiotensin-converting enzyme (ACE) inhibitor and a beta-blocker licensed to treat heart failure but only start one drug at a time.<br>For a person with confirmed heart failure with preserved ejection fraction:<br>- Ensure drugs which may cause or worsen heart failure are reviewed and stopped if appropriate.<br>- Prescribe a loop diuretic — up to 80 mg furosemide (or equivalent), if necessary, to relieve symptoms of fluid overload.<br>- Refer the person to a specialist for further advice on management.<br>- Consider if an antiplatelet drug is indicated.<br>- Consider prescribing an antiplatelet to people with atherosclerotic arterial disease (including coronary heart disease).<br>- Consider if statin therapy is indicated.<br>- Ensure that any causes, comorbidities, and precipitating factors are optimally managed.<br>- People with heart failure due to valve disease should be referred for specialist assessment and advice regarding follow-up. |

*Are there clear benefits of earlier detection or treatment?*

(more evidence or expert input needed)

#### Tests to screen for adverse treatment effects

- Treatments considered:
  - Statins
- Adverse treatment effects considered:
  - Those mentioned in BNF:
    - Common: myalgia and muscle pain; anaemia
    - Uncommon: hepatic disorders,
    - Rare or very rare: haematuria (renal related); polyneuropathy, myopathy
    - Frequency unknown: diabetes mellitus
- Corresponding lab tests considered:
  - Renal function tests
  - Liver function tests
  - Hba1c
  - Anaemia
  - Creatine kinase (for muscle symptoms)

#### Testing for Diabetes

*Is increased blood glucose or new onset diabetes more common in people who take antihypertensive drugs?*

Conflicting evidence on increased new-onset diabetes for statin users, with 3 studies suggesting no increased risk and 5 suggesting a slight increased risk (conflicting evidence). 3 of these studies suggest Rosuvastatin is specifically associated with increases risk of new onset diabetes.

| Source | Blood Tests | Evidence | Quality Concerns |
|---|---|---|---|
| BNF | Polyneuropathy was listed as rare (1 in 10,000 to 1 in 1,000) to very rare (less than 1 in 10,000) for statin users. Frequency was unknown for diabetes mellitus as a side effect. | | |
| *Systematic Reviews* | | | |
| Chou 2022 [20] | no specific blood tests mentioned | • Population: Adults 40 years or older without prior CVD events; studies of populations in which less than 10% of participants had prior CVD events were also eligible.<br>• Sample size: 26 studies (22 RCTs (N = 95 768, reported in 61 publications) assessed the effects of statins vs placebo or no statin; 1 RCT (n = 5144) of higher- vs lower-intensity statin therapy; and 3 large observational studies (n = 417 523)).<br>• Findings:<br>○ There was no significant association between statins and increased risk of (variably defined) incident diabetes (6 trials, n = 59 083 RR; 1.04 [95% CI, 0.92 to 1.19], although statistical heterogeneity was present.<br>○ JUPITER, the only trial to evaluate high-intensity Rosuvastatin statin therapy, was also the only trial to find increased risk (n = 17 802; 3.0% vs 2.4; RR, 1.25 [95%CI, 1.05 to 1.49]).<br>**Conclusion: high-intensity rosuvastatin statin therapy was found to increase risk of new onset diabetes, whereas no non- high-intensity statin therapy study found an increased risk.** | heterogeneity was present between many of the studies included for diabetes risk. |
| Oordt 2021 [21] | no specific blood tests mentioned | • Population: Adults without previous cardiovascular events<br>• Sample size: 18 studies on Statins licensed in Switzerland and at least one outcome, Placebo or no treatment and/or adaption for lifestyle (smoking reduction or stop, diet adaptation, physical activity))<br>• Findings:<br>○ No statistically significant difference between the statin and control groups in the occurrence of diabetes mellitus (RR: 1.04 , (95% CI 0.91-1.19).<br>**Conclusion: no significant risk of diabetes mellitus type 2 was found for statin users.** | studies reporting diabetes were low quality. |
| Singh 2020 [22] | no specific blood tests mentioned | • Population: participants without CVD with an average LDL-C between 100 and 159 mg<br>• Sample Size: 11 studies (n = 58504 (29235 in the statin group and 29269 controls)<br>• Findings:<br>○ the meta-analysis did not show any statistically significant difference between the statin and control groups in the occurrence of diabetes mellitus (RR 1.10, (95% CI:0.99 to 1.22)), | no |

| Source | Blood Tests | Evidence | Quality Concerns |
|---|---|---|---|
| | | **Conclusion: no significant risk of diabetes mellitus type 2 was found for statin users.** | |
| Zhao 2019 [23] | no specific blood tests mentioned | • Population: patients with hypercholesterolemia<br>• Sample size: 84 studies (n= 246,706). RCTs from phase 2 or higher evaluating statins, ezetimibe, and PCSK9 inhibitors monotherapy and comparing them against each other or placebo in adults with hypercholesterolemia.<br>• Findings:<br>○ Compared with placebo, statins were associated with increase of new-onset diabetes (1.13, [95% CI 1.02–1.26])<br>**Conclusion: Slight increase of new-onset diabetes in statin users.** | no |
| Rahal 2016 [24] | Fasting glucose | • Population: non-diabetics<br>• Sample size: 14 studies (n = 94,943)<br>• Findings:<br>○ The OR of diabetes incidence with statin therapy was significantly higher as compared with the placebo group (OR=1.11; 95% confidence interval = 1.0 to 1.2; p = 0.007).<br>○ Only 2 statins, atorvastatin (OR = 1.29 [95% CI 1.0-1.6; p = 0.042]) and rosuvastatin (OR = 1.17 [95% CI 1.0-1.3; p = 0.01]) were significantly associated with diabetes.<br>**Conclusion: significant increase of new-onset diabetes in atorvastatin and rosuvastatin users.** | No |
| Thakker 2016 [25] | Fasting glucose | • Population: general population<br>• Sample size: 29 studies (n= 163,039)<br>• Findings:<br>○ The meta-analysis derived that, statins significantly increased the likelihood of developing diabetes to 12% (pooled OR 1.12 [95%CI 1.05–1.21]; 18 RCTs) in the random-effects model. Only rosuvastatin was shown to increase the risk of developing diabetes significantly (pooled OR 1.18 [95%CI 1.04–1.33]; 4 RCTs).<br>**Conclusion: significant increase of new-onset diabetes rosuvastatin users.** | No |
| Vallejo-Vas 2015 [26] | Blood glucose and HbA1c | • Population: participants without diabetes at baseline,<br>• Sample size: 15 studies (4,815 non-diabetic patients (3,236 allocated to pitavastatin and 1,579 to control)<br>• Findings:<br>○ No significant differences associated with pitavastatin (vs. control) were observed for fasting blood glucose (mean difference -0.01 mg/dL [95% CI - 0.77-0.74]), HbA1c (mean difference -0.03% [95%CI 0.11-0.05]) or new onset diabetes (RR 0.70 [95% CI 0.30-1.61]).<br>**Conclusion: pitavastatin does not increase the risk of diabetes, or increase fasting glucose or HbA1c levels in users.** | Only reports for pitavastatin |
| Sattar 2010 [27] | Fasting blood glucose | • Population: general population, patients with organ transplants or receiving haemodialysis were excluded.<br>• Sample size: 13 studies (n = 91140) | No |

| Source | Blood Tests | Evidence | Quality Concerns |
|---|---|---|---|
| | | • Findings:<br>○ Statin therapy was associated with a 9% increased risk for incident diabetes (OR 1.09 [95% CI 1.02–1.17]), with little heterogeneity between trials.<br>**Conclusion: Statin therapy was associated with a slight increased risk for incident diabetes.** | |
| *Primary Studies* | | | |
| Search not needed. | | | |

*Is there anything the GP can do to manage or treat increased blood glucose or new onset diabetes?*

Yes, other than stopping or changing medication and dosage, there are several options for medications which can be changed in dose or combination to control blood glucose levels.

*Are there clear benefits of earlier detection or treatment?*

Yes, early intervention with drugs has long term benefits through lowering blood glucose and reducing the complications of Diabetes.

#### Renal function tests

*Is abnormal renal function more common in those who take hypertensive drugs?*

We found conflicting evidence to suggest statin users have a slight increase risk of renal insufficiency. One systematic review showed that Rosuvastatin was associated with increased risk of renal insufficiency.

| Source | Blood Tests | Evidence | Quality concerns |
|---|---|---|---|
| BNF | Haematuria (renal related) was listed as a rare (1 in 10,000 to 1 in 1,000) to very rare (less than 1 in 10,000) effect of statins. | | |
| *Systematic reviews* | | | |
| Cai 2021 [28] | proteinuria used, no specific blood test mentioned | • Population: Adults >18 years without a history of cardiovascular disease<br>• Sample size: 62 studies (n = 120,456), RCTs that compared statins with non-statin controls or compared different types or dosages of statins and reported at least one outcome of interest. 8 studies reported renal outcomes which included the presence of proteinuria (4 studies) and non-specified renal disorders (4 studies).<br>• Findings:<br>○ Statins were associated with renal insufficiency (8 studies, OR 1.14 [95% CI 1.01-1.28]).<br>○ Rosuvastatin specifically was associated with an increased risk of renal insufficiency (11 studies, OR 1.13 [95% CI 1.00-1.28]).<br>○ No other significant differences between the types of statins for renal results was found.<br>**Conclusion: Slight increase in renal insufficiency for statin users, this is mostly associated with Rosuvastatin.** | No |
| Oordt 2021 [21] | no specific blood test mentioned | • Population: Adults without previous cardiovascular events<br>• Sample size: 18 studies on statins licensed in Switzerland and at least one outcome, placebo or no treatment and/or adaption for lifestyle (smoking reduction or stop, diet adaptation, physical activity).<br>• Findings:<br>○ Slight increase risk in renal dysfunction (RR 1.12 [95% CI 1.00-1.26]).<br>**Conclusion: Slight increase in renal insufficiency for statin users.** | Studies reporting diabetes were low quality. |
| Taylor 2013 [29] | no specific blood test mentioned | • Population: adults with no restrictions on cholesterol levels, and where 10% or less had a history of CVD.<br>• Sample size: 18 Studies (n = 56,934) We included randomised controlled trials of statins versus placebo or usual care control with minimum treatment duration of one year and follow-up of six months,<br>• Findings:<br>○ No significant evidence was found for an increased risk of renal dysfunction (4 studies n = 27804, RR 1.11 [95% CI 0.99-1.26]).<br>**Conclusion: there is no significant increase in renal dysfunction in CKD patients that use statins.** | No |

| *Primary studies* |
| --- |
| Search not needed. |

*Is there anything the GP can do to manage or treat increased abnormal renal function?*

Other than stopping or changing statin and dosage and lifestyle changes there is not much that can be done.

*Are there clear benefits of earlier detection or treatment?*

Yes, early intervention can halt further kidney damage due to use of statins.

#### Liver function tests

*Is hepatitis or abnormal liver function more common in people who take antihypertensive drugs?*

Evidence suggests that statins increase the risk of liver injury, and raised alanine aminotransferase and aspartate aminotransferase levels. 1 study suggests lovastatin, 1 study suggests Fluvastatin, and 2 studies suggests Atorvastatin are specifically associated with the risk of liver injury.

| **Source** | **Blood Tests** | **Evidence** | **Quality Concerns** |
|---|---|---|---|
| BNF | Hepatic disorders are listed as uncommon (1 in 1000 to 1 in 100) in statin users. | | |
| *Systematic Reviews* | | | |
| Chou 2022 [20] | alanine aminotransferase and aspartate aminotransferase. | • Population: Adults 40 years or older without prior CVD events; studies of populations in which less than 10% of participants had prior CVD events were also eligible.<br>• Sample size: 26 studies (22 RCTs (N = 95,768, reported in 61 publications) assessed the effects of statins vs placebo or no statin; 1 RCT (n = 5,144) of higher- vs lower-intensity statin therapy; and 3 large observational studies (n = 417,523)).<br>• Findings: Statin therapy, vs placebo or no statin, was not significantly associated with<br>○ elevated alanine aminotransferase level (10 trials (n = 48,149); RR 0.94 [95% CI 0.78-1.13]).<br>○ or elevated aspartate aminotransferase level (4 trials (n = 17,534); RR 1.30 [95% CI 0.78 - 2.17]).<br>**Conclusion: Statins do not increase the risk of elevated alanine aminotransferase and elevated aspartate aminotransferase levels.** | No |
| Cai 2021 [28] | aspartate transaminase or alanine Transaminase | • Population: Adults >18 years without a history of cardiovascular disease<br>• Sample size: 62 studies (n = 120,456), RCTs that compared statins with non-statin controls or compared different types or dosages of statins and reported at least one outcome of interest.<br>• Findings:<br>○ Statins increased the risk of liver dysfunction (21 studies, odds ratio 1.33 [95% CI 1.12 - 1.58]); which was defined as raised serum concentration of liver enzymes (aspartate transaminase or alanine<br>○ transaminase) in all studies.<br>○ Atorvastatin (17 studies, OR 1.41 [955 CI 1.08 - 1.85]) and lovastatin (five studies, OR 1.81 [95% CI 1.23 - 2.66]) increased the risk of liver dysfunction.<br>**Conclusion: Statins increase the risk of liver dysfunction. Sub-analysis shows lovastatin and atorvastatin specifically are associated with increased liver dysfunction.** | No |
| Singh 2020 [22] | AST/ALT elevation | • Population: participants free of CVD with an average LDL-C between 100 and 159 mg.<br>• Sample size: 11 studies (n = 58,504 (29,235 in the statin group and 29,269 controls).<br>• Findings:<br>○ Weak evidence was found for an increased risk of liver enzyme abnormalities: RR 1.20 [95% CI 1.00-1.43]). | No |

| Source | Blood Tests | Evidence | Quality Concerns |
|---|---|---|---|
| | | **Conclusion: Statins slightly increased risk of liver enzyme abnormalities.** | |
| Zhao 2019 [23] | ALT levels | • Population: patients with hypercholesterolemia<br>• Sample Size: 84 studies (n= 246,706). RCTs from phase 2 or higher evaluating statins, ezetimibe, and PCSK9 inhibitors monotherapy were compared against each other or placebo in adults with hypercholesterolemia.<br>• Findings:<br>○ Compared with placebo, statins were associated with elevation of ALT (OR 1.89 [95% CI 1.42–2.51]).<br>**Conclusion: Statins slightly increased risk of raised alanine transaminase levels.** | No |
| Liang 2018 [30] | Liver injury | • Population: adults older than 18 years old<br>• Sample size: 16 studies (n = 74, 078) (RCTs design as either parallel or crossover; investigating the influence of statins on liver function).<br>• Findings:<br>○ The pooled odds ratio of 16 studies was 1.18 [95% CI 1.01-1.39], indicating that statin therapy could increase the liver injury.<br>○ Subgroup analysis indicated that Fluvastatin increased the risk of liver injury significantly (OR: 3.50 [95% CI 1.07–11.53]) and dose over 40mg/daily had an unfavourable effect on the liver damage (OR: 3.62 [95% CI 1.52–8.65]).<br>**Conclusion: Statins slightly increase the risk of liver injury, with Fluvastatin and dosages above 40mg per day increase in the risk of liver injury significantly.** | No |
| Li 2016 [31] | Liver function tests | • Population: healthy adults or adults with, or at risk of developing, cardiovascular or cerebrovascular disease<br>• Sample size: 17 studies (n = 21,910 (68.8 % male)): n= 11,107 for the atorvastatin 80 mg/day group and n = 10,803 for the control group.<br>• Findings:<br>○ the atorvastatin 80 mg/day group had a much higher risk of ALT/AST elevation (RR 4.59 [95 % CI 3.26–6.48]) compared with controls.<br>○ Short-term treatment with atorvastatin 80 mg/day did not increase the incidence of transaminase elevation (RR 2.05 [95 % CI 0.64–6.53]), but long-term treatment did (RR 4.90 [95 % CI 3.42–7.04]).<br>**Conclusion: Atorvastatin significantly increases the risk of raised ALT/AST levels, but only during long-term use.** | No |
| *Primary Studies* | | | |
| Search not needed. | | | |

*Is there anything the GP can do to manage or treat increased abnormal liver function?*

Other than stopping or changing medication and dosage and lifestyle changes there is not much that can be done.

*Are there clear benefits of earlier detection or treatment?*

Yes, early intervention can halt further liver damage by statins.

#### Anaemia

*Is Anaemia more common in people who take antihypertensive drugs?*

Evidence suggests that Anaemia is not a side effect of Statins except for an increased risk of Vitamin B12 deficiency in men.

| Source | Blood Tests | Evidence | Quality Concerns |
|---|---|---|---|
| BNF | Anaemia was listed as a common (1 in 100 to 1 in 10) side effect for statins. | | |
| *Systematic Reviews* | | | |
| None identified | | | |
| *Primary Studies* | | | |
| Masajtis-Zagajewska 2018 [32] | Haemoglobiln (Hb), Serum Ferritin and Serum folate | • Population: Adults with stable CKD for 6 months and without anaemia<br>• Sample size: RCT of 36 patients (19 women and 17 men)<br>• Findings:<br>○ Change in Hemoglobin (g/dL) statin vs placebo: 0.06 ± 0.9 [95% CI –0.24-0.36]<br>○ Change in Iron(g/dL) statin vs placebo: 0 ± 9.2 [95% CI –3.09-3.09]<br>○ Change in Ferritin (ng/mL) statin vs placebo: –0.68 ± 64.3 [95% CI 22.3–20.9]<br>**Conclusion: No significant change in haemoglobin, iron or ferritin levels was seen when on statins.** | Small sample size and area sampled, unlikely to be representative of the population. |
| Bolaman 2006 [33] | Haemoglobiln (Hb), Vitamin B12, folic acid | • Population: patients with primary hypercholesterolemia<br>• Sample size: RCT of 44 patients (37 women, 7 men)<br>• Findings:<br>○ Vitamin-B 12 levels significantly increased after atorvastatin treatment, but other hematological parameters did not change. This increase was significant only in male patients.<br>**Conclusion: Increase in vitamin B12 after starting statins in men, no other significant change seen.** | Small sample size and area sampled, sample size of just men even smaller, unlikely to be representative of the population |

*Is there anything the GP can do to manage or treat Anaemia?*

Yes, give dietary advice and for iron deficiency anaemia prescribe iron tablets, for Vit B12 deficiency anaemia refer or administer hydroxocobalamin, for folate deficiency anaemia prescribe folic acid.

*Are there clear benefits of earlier detection or treatment?*

Yes, early intervention can halt further lowering of iron, folate, vitamin B12, and haemoglobin levels.

#### Creatine Kinase

*Are muscle symptoms and disorders more common in people who take Statins?*

No, evidence suggests that people taking statins have no increased risk of myalgia, muscle pain, or rhabdomyolysis. However there is conflicting evidence on the risk of increased creatine kinase, with one study reporting a slight risk of increased Creatine Kinase levels and 2 reporting no increased levels.

| **Source** | **Blood test** | **Evidence** | **Quality concerns** |
|---|---|---|---|
| BNF | Myalgia and muscle pain was listed as a common (1 in 100 to 1 in 10) side effect for statins. Myopathy was listed as rare (1 in 10,000 to 1 in 1,000) to very rare (less than 1 in 10,000). | | |
| *Systematic reviews* | | | |
| Chou 2022 [20] | Creatine Kinase | • Population: Adults 40 years or older without prior CVD events; studies of populations in which less than 10% of participants had prior CVD events were also eligible.<br>• Sample size: 26 studies (22 RCTs (N = 95,768, reported in 61 publications) assessed the effects of statins vs placebo or no statin; 1 RCT (n = 5,144) of higher- vs lower-intensity statin therapy; and 3 large observational studies (n = 417,523)).<br>• Findings:<br>o Statin therapy, vs placebo or no statin, was not significantly associated with myalgia (9 trials, n = 46,388; RR: 0.98 [95% CI 0.86-1.11]).<br>o Statins were also not significantly associated with increased risk of myopathy (3 trials, n = 33,345; RR: 1.09 [95%CI 0.48-2.47]).<br>o or rhabdomyolysis (4 trials, n = 59,672; RR: 1.54 [95% CI 0.36-6.64]).<br>**Conclusion: Statins do not increase risk of myalgia, myopathy or rhabdomyolysis.** | No |
| Singh 2020 [22] | Creatine kinase (CK) | • Population: participants free of CVD with an average LDL-C between 100 and 159 mg.<br>• Sample Size: 11 studies (n = 58,504 (29,235 in the statin group and 29,269 controls).<br>• Findings:<br>o Myalgia: RR 1.02 [95% CI 0.88-1.19].<br>o Creatine kinase elevation: RR 0.88 [95% CI 0.51-1.53].<br>o Rhabdomyolysis: RR 1.29 [95% CI 0.25-6.69].<br>o Myopathy: RR 1.09 [95% CI 0.48-2.47].<br>**Conclusions: no significant increased risk was seen for adverse muscle symptoms in statin users.** | No |
| Zhao 2019 [23] | Creatine Kinase (CK) | • Population: patients with hypercholesterolemia | No |

| Source | Blood test | Evidence | Quality concerns |
|---|---|---|---|
|  |  | • Sample Size: 84 studies (n= 246,706). RCTs from phase 2 or higher evaluating statins, ezetimibe, and PCSK9 inhibitors monotherapy and comparing them against each other or placebo in adults with hypercholesterolemia.<br>• Findings:<br> ○ Compared with placebo, statins were associated with elevation of CK levels (OR 1.45 [95% CI 1.09–1.93].<br>**Conclusion: Statin users have a slight risk of increased creatine kinase levels.** |  |
| Li 2016 [31] | Creatine Kinase (CK) | • Population: healthy adults or adults with, or at risk of developing, cardiovascular or cerebrovascular disease<br>• Sample size: 17 studies (n = 21,910 (68.8 % male)): n= 11,107 for the atorvastatin 80 mg/day group and n = 10,803 for the control group. Information regarding CK elevation was available in 11 studies involving 7,438 participants.<br>• Findings:<br> ○ No significant difference was observed between the two groups (RR 1.38 [95% CI 0.97–1.95]).<br> ○ The pooled analysis suggested that atorvastatin 80 mg/day was similar to controls in terms of myalgia incidence (RR 1.06 [95% CI 0.93–1.20]).<br> ○ The meta-analysis demonstrated that atorvastatin 80 mg/day did not increase the incidence of rhabdomyolysis (RR 0.67 [95 %CI 0.19–2.36]).<br> ○ No significant difference was observed in CK elevation between the two groups (RR 1.38 [95% CI 0.97–1.95]).<br>**Conclusion: no significant increased risk was seen for adverse muscle symptoms or increased CK levels in statin users.** | No |
| *Primary studies* | | | |
| Search not needed. | | | |

*Is there anything the GP can do to manage or treat these side effects?*

Other than stopping or changing medication and dosage and lifestyle changes there is not much that can be done.

*Are there clear benefits of earlier detection or treatment?*

Yes, early intervention can halt further raising of creatine levels, if statins are the cause.

#### Abbreviations

| | |
|---|---|
| ACE | Angiotensin-converting enzyme |
| ALP | Alkaline phosphatase |
| ALT | Alanine aminotransferase |
| AST | Aspartate transaminase |
| BNF | British National Formulary |
| BNP | Brain Natriuretic Peptide |
| CGM | Continuous Glucose Monitoring |
| CKD | Chronic Kidney Disease |
| CRP | C-reactive protein |
| CVD | Cardiovascular disease |
| eGFR | Estimated Glomerular Filtration Rate |
| ESR | Erythrocyte Sedimentation Rate |
| ESRD | End-stage renal disease |
| FPG | Fasting plasma glucose |
| GA | Glycated albumin |
| HbA1c | Glycated haemoglobin |
| HDL | High-density lipoprotein |
| HF | Heart failure |
| HTN | Hypertension |
| ICHOM | International Consortium for Health Outcomes Measurement |
| KNHANES | Korean National Health and Nutrition Examination Survey |
| LDL | Low-density lipoprotein |
| MI | Myocardial infarction |
| NAFLD | Non-alcoholic fatty liver disease |
| NHANES | National Health and Nutrition Examination Survey |
| NICE | National Institute for Health and Care Excellence |
| PVD | Peripheral Vascular Disease |
| RRT | Renal Replacement Therapy |
| T1DM | Type 1 diabetes mellitus |
| T2DM | Type 2 diabetes mellitus |
| UKPDS | UK Prospective Diabetes Study |

#### Table S11: Bone profile codes

| Code | Description | Coding system | Concept name | Phenotype name |
|---|---|---|---|---|
| Cyu4100 | [X]Other hyperparathyroidism | Read codes v2 | Hyperparathyroidism - Primary care | Hyperparathyroidism |
| C120000 | Primary hyperparathyroidism | Read codes v2 | Hyperparathyroidism - Primary care | Hyperparathyroidism |
| C120200 | Tertiary hyperparathyroidism | Read codes v2 | Hyperparathyroidism - Primary care | Hyperparathyroidism |
| C120111 | Osteitis fibrosa cystica | Read codes v2 | Hyperparathyroidism - Primary care | Hyperparathyroidism |
| K08y100 | Secondary hyperparathyroidism | Read codes v2 | Hyperparathyroidism - Primary care | Hyperparathyroidism |
| C120.00 | Hyperparathyroidism | Read codes v2 | Hyperparathyroidism - Primary care | Hyperparathyroidism |
| N332500 | Brown tumour of hyperparathyroidism | Read codes v2 | Hyperparathyroidism - Primary care | Hyperparathyroidism |
| C120100 | Hyperparathyroid bone disease | Read codes v2 | Hyperparathyroidism - Primary care | Hyperparathyroidism |
| C120.12 | Von Recklinghausen's bone disease | Read codes v2 | Hyperparathyroidism - Primary care | Hyperparathyroidism |
| C1z3100 | Ectopic hyperparathyroidism | Read codes v2 | Hyperparathyroidism - Primary care | Hyperparathyroidism |
| C120112 | Von Recklinghausen's bone disease | Read codes v2 | Hyperparathyroidism - Primary care | Hyperparathyroidism |
| C120.11 | Osteitis fibrosa cystica | Read codes v2 | Hyperparathyroidism - Primary care | Hyperparathyroidism |
| 19457 | Diagnosis of Hyperparathyroidism | Med codes | Hyperparathyroidism - Primary care | Hyperparathyroidism |
| 3559 | Diagnosis of Hyperparathyroidism | Med codes | Hyperparathyroidism - Primary care | Hyperparathyroidism |
| 56524 | Diagnosis of Hyperparathyroidism | Med codes | Hyperparathyroidism - Primary care | Hyperparathyroidism |
| 105537 | Diagnosis of Hyperparathyroidism | Med codes | Hyperparathyroidism - Primary care | Hyperparathyroidism |
| 73934 | Diagnosis of Hyperparathyroidism | Med codes | Hyperparathyroidism - Primary care | Hyperparathyroidism |
| 4116 | Diagnosis of Hyperparathyroidism | Med codes | Hyperparathyroidism - Primary care | Hyperparathyroidism |
| 17339 | Diagnosis of Hyperparathyroidism | Med codes | Hyperparathyroidism - Primary care | Hyperparathyroidism |
| 49456 | Diagnosis of Hyperparathyroidism | Med codes | Hyperparathyroidism - Primary care | Hyperparathyroidism |
| 53819 | Diagnosis of Hyperparathyroidism | Med codes | Hyperparathyroidism - Primary care | Hyperparathyroidism |
| 65478 | Diagnosis of Hyperparathyroidism | Med codes | Hyperparathyroidism - Primary care | Hyperparathyroidism |
| 73923 | Diagnosis of Hyperparathyroidism | Med codes | Hyperparathyroidism - Primary care | Hyperparathyroidism |
| 18740 | Diagnosis of Hyperparathyroidism | Med codes | Hyperparathyroidism - Primary care | Hyperparathyroidism |
| E21.1 | Secondary hyperparathyroidism, not elsewhere classified | ICD10 codes | Hyperparathyroidism - Secondary care - Diagnoses | Hyperparathyroidism |

| | | | | |
|---|---|---|---|---|
| E21.0 | Primary hyperparathyroidism | ICD10 codes | Hyperparathyroidism - Secondary care - Diagnoses | Hyperparathyroidism |
| E21.3 | Hyperparathyroidism, unspecified | ICD10 codes | Hyperparathyroidism - Secondary care - Diagnoses | Hyperparathyroidism |
| E21.2 | Other hyperparathyroidism | ICD10 codes | Hyperparathyroidism - Secondary care - Diagnoses | Hyperparathyroidism |
| N331N00 | Fragility fracture | Read codes v2 | Osteoporosis - Primary care | Osteoporosis |
| N331B00 | Postmenopausal osteoporosis with pathological fracture | Read codes v2 | Osteoporosis - Primary care | Osteoporosis |
| N331200 | Postoophorectomy osteoporosis with pathological fracture | Read codes v2 | Osteoporosis - Primary care | Osteoporosis |
| N330D00 | Osteoporosis due to corticosteroids | Read codes v2 | Osteoporosis - Primary care | Osteoporosis |
| N330400 | Dissuse osteoporosis | Read codes v2 | Osteoporosis - Primary care | Osteoporosis |
| N330.00 | Osteoporosis | Read codes v2 | Osteoporosis - Primary care | Osteoporosis |
| 9Od0.00 | Attends osteoporosis monitoring | Read codes v2 | Osteoporosis - Primary care | Osteoporosis |
| 58EG.00 | Hip DXA scan result osteoporotic | Read codes v2 | Osteoporosis - Primary care | Osteoporosis |
| NyuB200 | [X]Osteoporosis in other disorders classified elsewhere | Read codes v2 | Osteoporosis - Primary care | Osteoporosis |
| 9Od6.00 | Osteoporosis monitoring verbal invitation | Read codes v2 | Osteoporosis - Primary care | Osteoporosis |
| N330700 | Postsurgical malabsorption osteoporosis | Read codes v2 | Osteoporosis - Primary care | Osteoporosis |
| N331A00 | Osteoporosis + pathological fracture cervical vertebrae | Read codes v2 | Osteoporosis - Primary care | Osteoporosis |
| 9Od9.00 | Osteoporosis monitoring check done | Read codes v2 | Osteoporosis - Primary care | Osteoporosis |
| N331600 | Idiopathic osteoporosis with pathological fracture | Read codes v2 | Osteoporosis - Primary care | Osteoporosis |
| N330600 | Postoophorectomy osteoporosis | Read codes v2 | Osteoporosis - Primary care | Osteoporosis |
| N330100 | Senile osteoporosis | Read codes v2 | Osteoporosis - Primary care | Osteoporosis |
| N331L00 | Collapse of vertebra due to osteoporosis NOS | Read codes v2 | Osteoporosis - Primary care | Osteoporosis |
| N331K00 | Collapse of thoracic vertebra due to osteoporosis | Read codes v2 | Osteoporosis - Primary care | Osteoporosis |
| N331M11 | Minimal trauma fracture due to unspecified osteoporosis | Read codes v2 | Osteoporosis - Primary care | Osteoporosis |
| N331M00 | Fragility fracture due to unspecified osteoporosis | Read codes v2 | Osteoporosis - Primary care | Osteoporosis |

| | | | | |
|---|---|---|---|---|
| N330A00 | Osteoporosis in endocrine disorders | Read codes v2 | Osteoporosis - Primary care | Osteoporosis |
| 9kj..00 | Osteoporosis - enhanced services administration | Read codes v2 | Osteoporosis - Primary care | Osteoporosis |
| N330200 | Postmenopausal osteoporosis | Read codes v2 | Osteoporosis - Primary care | Osteoporosis |
| 9Od2.00 | Osteoporosis monitoring default | Read codes v2 | Osteoporosis - Primary care | Osteoporosis |
| NyuB800 | [X]Unspecified osteoporosis with pathological fracture | Read codes v2 | Osteoporosis - Primary care | Osteoporosis |
| NyuB100 | [X]Other osteoporosis | Read codes v2 | Osteoporosis - Primary care | Osteoporosis |
| N331H00 | Collapse of cervical vertebra due to osteoporosis | Read codes v2 | Osteoporosis - Primary care | Osteoporosis |
| 9Od8.00 | Osteoporosis monitoring deleted | Read codes v2 | Osteoporosis - Primary care | Osteoporosis |
| N330C00 | Osteoporosis localized to spine | Read codes v2 | Osteoporosis - Primary care | Osteoporosis |
| 9Od..00 | Osteoporosis monitoring administration | Read codes v2 | Osteoporosis - Primary care | Osteoporosis |
| N330500 | Drug-induced osteoporosis | Read codes v2 | Osteoporosis - Primary care | Osteoporosis |
| N330z00 | Osteoporosis NOS | Read codes v2 | Osteoporosis - Primary care | Osteoporosis |
| NyuB000 | [X]Other osteoporosis with pathological fracture | Read codes v2 | Osteoporosis - Primary care | Osteoporosis |
| 9Od3.00 | Osteoporosis monitoring first letter | Read codes v2 | Osteoporosis - Primary care | Osteoporosis |
| 58EM.00 | Lumbar DXA scan result osteoporotic | Read codes v2 | Osteoporosis - Primary care | Osteoporosis |
| N331800 | Osteoporosis + pathological fracture lumbar vertebrae | Read codes v2 | Osteoporosis - Primary care | Osteoporosis |
| N374600 | Osteoporotic kyphosis | Read codes v2 | Osteoporosis - Primary care | Osteoporosis |
| N331300 | Osteoporosis of disuse with pathological fracture | Read codes v2 | Osteoporosis - Primary care | Osteoporosis |
| 9Od5.00 | Osteoporosis monitoring third letter | Read codes v2 | Osteoporosis - Primary care | Osteoporosis |
| N330B00 | Vertebral osteoporosis | Read codes v2 | Osteoporosis - Primary care | Osteoporosis |
| 66a..00 | Osteoporosis monitoring | Read codes v2 | Osteoporosis - Primary care | Osteoporosis |
| 9Od7.00 | Osteoporosis monitoring telephone invitation | Read codes v2 | Osteoporosis - Primary care | Osteoporosis |
| N331900 | Osteoporosis + pathological fracture thoracic vertebrae | Read codes v2 | Osteoporosis - Primary care | Osteoporosis |
| N331J00 | Collapse of lumbar vertebra due to osteoporosis | Read codes v2 | Osteoporosis - Primary care | Osteoporosis |
| N331400 | Postsurgical malabsorption | Read codes v2 | Osteoporosis - Primary care | Osteoporosis |

| | | | | |
|---|---|---|---|---|
| | osteoporosis with path fracture | | | |
| 14GB.00 | History of osteoporosis | Read codes v2 | Osteoporosis - Primary care | Osteoporosis |
| 9kj0.00 | Bone sparing drug treatment offered for osteoporosis - ESA | Read codes v2 | Osteoporosis - Primary care | Osteoporosis |
| N330300 | Idiopathic osteoporosis | Read codes v2 | Osteoporosis - Primary care | Osteoporosis |
| N330000 | Osteoporosis, unspecified | Read codes v2 | Osteoporosis - Primary care | Osteoporosis |
| 9Od4.00 | Osteoporosis monitoring second letter | Read codes v2 | Osteoporosis - Primary care | Osteoporosis |
| N331N11 | Minimal trauma fracture | Read codes v2 | Osteoporosis - Primary care | Osteoporosis |
| N331500 | Drug-induced osteoporosis with pathological fracture | Read codes v2 | Osteoporosis - Primary care | Osteoporosis |
| 58EV.00 | Femoral neck DEXA scan result osteoporotic | Read codes v2 | Osteoporosis - Primary care | Osteoporosis |
| 45736 | Diagnosis of Osteoporosis | Med codes | Osteoporosis - Primary care | Osteoporosis |
| 57301 | Diagnosis of Osteoporosis | Med codes | Osteoporosis - Primary care | Osteoporosis |
| 40428 | Diagnosis of Osteoporosis | Med codes | Osteoporosis - Primary care | Osteoporosis |
| 33526 | Diagnosis of Osteoporosis | Med codes | Osteoporosis - Primary care | Osteoporosis |
| 16857 | Diagnosis of Osteoporosis | Med codes | Osteoporosis - Primary care | Osteoporosis |
| 42354 | Diagnosis of Osteoporosis | Med codes | Osteoporosis - Primary care | Osteoporosis |
| 62702 | Diagnosis of Osteoporosis | Med codes | Osteoporosis - Primary care | Osteoporosis |
| 39334 | Diagnosis of Osteoporosis | Med codes | Osteoporosis - Primary care | Osteoporosis |
| 36796 | History of Osteoporosis | Med codes | Osteoporosis - Primary care | Osteoporosis |
| 48772 | Diagnosis of Osteoporosis | Med codes | Osteoporosis - Primary care | Osteoporosis |
| 25534 | History of Osteoporosis | Med codes | Osteoporosis - Primary care | Osteoporosis |
| 96779 | History of Osteoporosis | Med codes | Osteoporosis - Primary care | Osteoporosis |
| 31580 | Diagnosis of Osteoporosis | Med codes | Osteoporosis - Primary care | Osteoporosis |
| 96342 | Diagnosis of Osteoporosis | Med codes | Osteoporosis - Primary care | Osteoporosis |
| 39217 | Diagnosis of Osteoporosis | Med codes | Osteoporosis - Primary care | Osteoporosis |
| 27597 | Diagnosis of Osteoporosis | Med codes | Osteoporosis - Primary care | Osteoporosis |
| 68019 | Diagnosis of Osteoporosis | Med codes | Osteoporosis - Primary care | Osteoporosis |
| 105290 | History of Osteoporosis | Med codes | Osteoporosis - Primary care | Osteoporosis |

| | | | | |
|---|---|---|---|---|
| 102730 | Diagnosis of Osteoporosis | Med codes | Osteoporosis - Primary care | Osteoporosis |
| 93981 | Diagnosis of Osteoporosis | Med codes | Osteoporosis - Primary care | Osteoporosis |
| 4013 | Diagnosis of Osteoporosis | Med codes | Osteoporosis - Primary care | Osteoporosis |
| 93497 | Diagnosis of Osteoporosis | Med codes | Osteoporosis - Primary care | Osteoporosis |
| 5841 | Diagnosis of Osteoporosis | Med codes | Osteoporosis - Primary care | Osteoporosis |
| 277 | Diagnosis of Osteoporosis | Med codes | Osteoporosis - Primary care | Osteoporosis |
| 16307 | Diagnosis of Osteoporosis | Med codes | Osteoporosis - Primary care | Osteoporosis |
| 98433 | History of Osteoporosis | Med codes | Osteoporosis - Primary care | Osteoporosis |
| 3346 | Diagnosis of Osteoporosis | Med codes | Osteoporosis - Primary care | Osteoporosis |
| 34798 | Diagnosis of Osteoporosis | Med codes | Osteoporosis - Primary care | Osteoporosis |
| 61121 | History of Osteoporosis | Med codes | Osteoporosis - Primary care | Osteoporosis |
| 41755 | Diagnosis of Osteoporosis | Med codes | Osteoporosis - Primary care | Osteoporosis |
| 93455 | History of Osteoporosis | Med codes | Osteoporosis - Primary care | Osteoporosis |
| 98760 | History of Osteoporosis | Med codes | Osteoporosis - Primary care | Osteoporosis |
| 38395 | Diagnosis of Osteoporosis | Med codes | Osteoporosis - Primary care | Osteoporosis |
| 19048 | Diagnosis of Osteoporosis | Med codes | Osteoporosis - Primary care | Osteoporosis |
| 9700 | Diagnosis of Osteoporosis | Med codes | Osteoporosis - Primary care | Osteoporosis |
| 11503 | Diagnosis of Osteoporosis | Med codes | Osteoporosis - Primary care | Osteoporosis |
| 68122 | History of Osteoporosis | Med codes | Osteoporosis - Primary care | Osteoporosis |
| 92887 | History of Osteoporosis | Med codes | Osteoporosis - Primary care | Osteoporosis |
| 12673 | Diagnosis of Osteoporosis | Med codes | Osteoporosis - Primary care | Osteoporosis |
| 25650 | Diagnosis of Osteoporosis | Med codes | Osteoporosis - Primary care | Osteoporosis |
| 36432 | Diagnosis of Osteoporosis | Med codes | Osteoporosis - Primary care | Osteoporosis |
| 102017 | History of Osteoporosis | Med codes | Osteoporosis - Primary care | Osteoporosis |
| 99817 | History of Osteoporosis | Med codes | Osteoporosis - Primary care | Osteoporosis |
| 93705 | Diagnosis of Osteoporosis | Med codes | Osteoporosis - Primary care | Osteoporosis |
| 24093 | Diagnosis of Osteoporosis | Med codes | Osteoporosis - Primary care | Osteoporosis |

| | | | | |
|---|---|---|---|---|
| 18825 | Diagnosis of Osteoporosis | Med codes | Osteoporosis - Primary care | Osteoporosis |
| 93655 | Diagnosis of Osteoporosis | Med codes | Osteoporosis - Primary care | Osteoporosis |
| 70349 | Diagnosis of Osteoporosis | Med codes | Osteoporosis - Primary care | Osteoporosis |
| 65163 | History of Osteoporosis | Med codes | Osteoporosis - Primary care | Osteoporosis |
| 17377 | Diagnosis of Osteoporosis | Med codes | Osteoporosis - Primary care | Osteoporosis |
| 46894 | Diagnosis of Osteoporosis | Med codes | Osteoporosis - Primary care | Osteoporosis |
| 11603 | History of Osteoporosis | Med codes | Osteoporosis - Primary care | Osteoporosis |
| 14967 | Diagnosis of Osteoporosis | Med codes | Osteoporosis - Primary care | Osteoporosis |
| M82 | Osteoporosis in diseases classified elsewhere | ICD10 codes | Osteoporosis - Secondary care - Diagnoses | Osteoporosis |
| M81 | Osteoporosis without pathological fracture | ICD10 codes | Osteoporosis - Secondary care - Diagnoses | Osteoporosis |
| M80 | Osteoporosis with pathological fracture | ICD10 codes | Osteoporosis - Secondary care - Diagnoses | Osteoporosis |
